# Environmental Screening for Surface SARS-CoV-2 Contamination in Urban High-Touch Areas

**DOI:** 10.1101/2021.05.04.21256107

**Authors:** Lauren Roppolo Brazell, Shawna Stetz, Adam Hipp, Samantha Taylor, Nicholas Stark, Katherine Jensen, Md Ariful Islam Juel, Patrick Deegan, Mariya Munir, Jessica Schlueter, Jennifer Weller, Cynthia Gibas

## Abstract

The novel human coronavirus 2 (SARS-CoV-2) is responsible for the COVID-19 outbreak, which reached pandemic-level infection rates in just a few short months after being identified in late 2019. Early transmission models focused on surface contamination, but current research provides evidence for person-to-person transmission via aerosolized viral particles. As such, the CDC’s guidance has recently been updated to increasingly redirect the focus of prevention methods to aerosol routes. Inhalation of SARS-CoV-2 particles presents the most significant threat of infection to individuals. A secondary route, from hand to mouth, eyes or nose, is likely after contact with a surface contaminated with particles that have settled out of aerosols or been deposited by contaminated hands. Using common molecular detection methods including endpoint and quantitative PCR, we investigated whether there is detectable contamination by SARS-CoV-2 on high-touch surfaces on public transit vehicles and on other high-touch surfaces on a college campus during normal use. Our results indicate that SARS-CoV-2 can be successfully recovered and detected on common high-touch surfaces, albeit in comparatively lower frequencies as public health guidance progressed and more rigorous sanitization procedures were implemented.

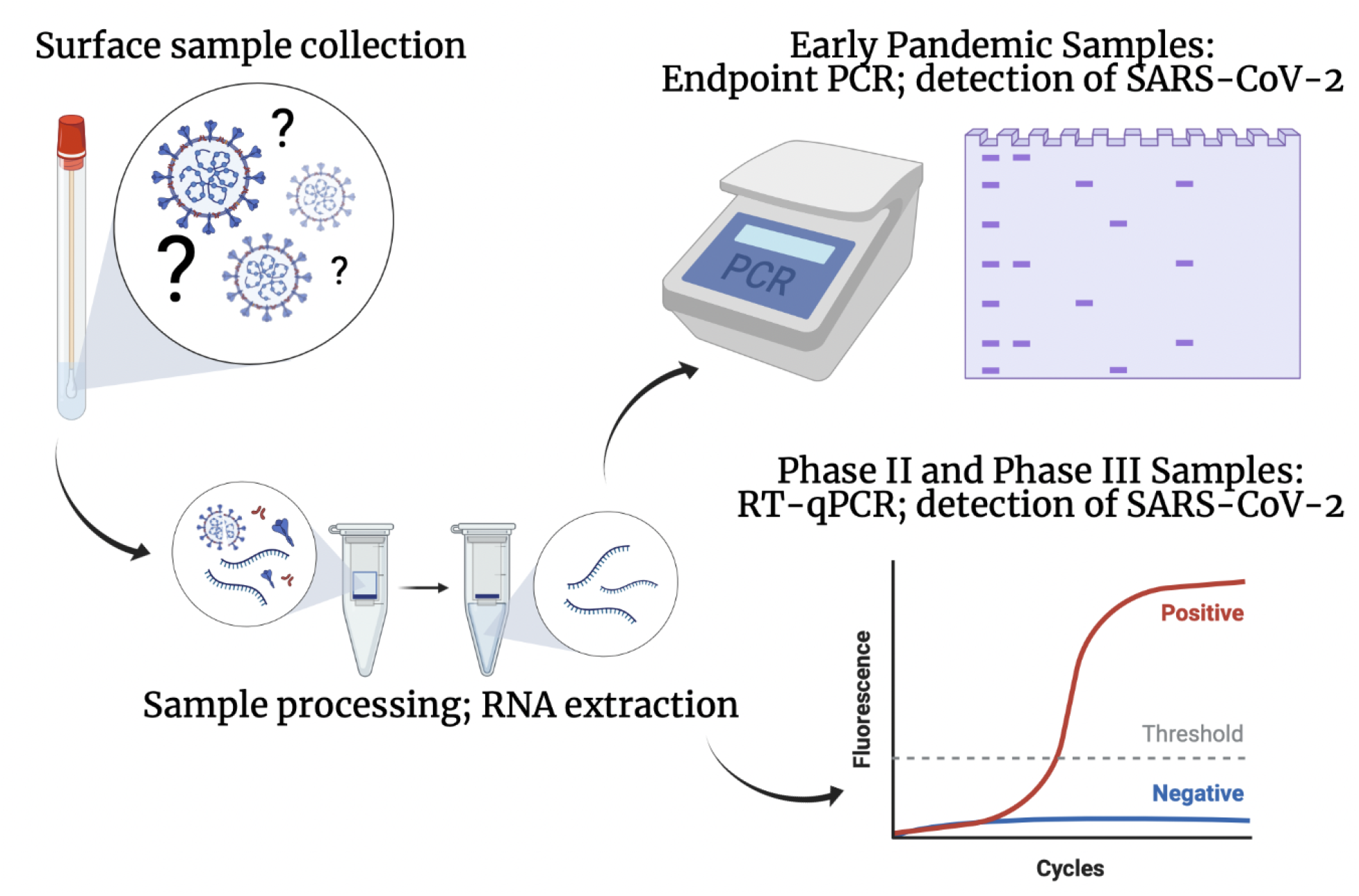

**Graphical abstract created with permission from BioRender.com (2021)**.

## Introduction

It has been over a year since the initial discovery of the Novel Human Coronavirus 2 (SARS-CoV-2), the virus responsible for the ongoing COVID-19 pandemic. We now know that our best defense against the virus is the CDC’s recommendations of mask wearing in public, maintaining safe social distance from others, and practicing good personal hygiene via handwashing [1]. Even with these precautions in place, the disease continues to spread rapidly. The World Health Organization’s most recently published data states that over 100 million cases have been reported, with over 2 million deaths reported worldwide [2].

While some individuals who fall ill with COVID-19 show symptoms with varying degrees of severity, others remain asymptomatic and continue their daily lives as normal, shedding the virus and infecting those that they come into close contact with [3]. When an infected individual coughs, sneezes or exhales, virus-containing particles are aerosolized. Even though the primary threat of transmission is the inhalation of these aerosolized viral particles [4], they can also infect individuals by way of contact with surfaces they settle on [3].

To better understand the surface transmissibility of COVID-19, many studies have been conducted to assay the presence of SARS-CoV-2 RNA on surfaces. Most of these studies have been conducted in clinical settings with high volumes of COVID-19 patients and inherent risk, revealing the presence of the virus on surfaces like stainless steel, polyester, and porous PPE [5]. A study conducted early in 2020 found that settled viral SARS-CoV-2 particles were viable on polyester and stainless steel surfaces for up to 72 hours after exposure [6]. Several months later, a more thorough study isolated viral RNA, inoculated different types of porous and non-porous high-touch surfaces and concluded that, under controlled conditions, viral particles could still be detected after 28 days on non-porous surfaces kept at 20° and 50% humidity [7].

With this in mind, we sought to design a study to assess the risk of encountering SARS-CoV-2 RNA on surfaces in high-traffic, high-touch areas within the Charlotte, North Carolina community. Charlotte is the most densely populous city in the state, with a 2019 US Census Bureau reported population of 885,708 [8]. We aimed to sample some of the busiest places in the city, including high-traffic buildings and buses on the local university campus, and vehicles in the local public transit system, which remained in operation during pandemic closures.

Prior to the adoption of any formal restrictions, local schools and universities were forced to make difficult, but potentially life-saving decisions regarding instruction and campus life. Officials at the University of North Carolina at Charlotte monitored the spread of the pandemic closely and ultimately chose to move to virtual instruction beginning on March 16 [9]. Students were then required to vacate their residence halls no later than March 20, unless there was an extenuating circumstance that prevented them from leaving. Services deemed essential were allowed to remain open during this time [10]. At the same time, the City of Charlotte began implementing a multiphase model for closing and reopening heavily populated areas in response to mounting COVID-19 infection rates (including businesses, schools, restaurants, etc.) to help reduce instances of close person-to-person contact that could potentially spread the virus. Mandated by the Governor of North Carolina, the state enacted a “Stay at Home Order” on May 8, 2020 (Executive Order 138), and remained under these orders until entering Phase II of restrictions on May 23, 2020. During Phase II, some restrictions were lifted on activities like in-person dining, social gatherings outdoors for sporting events and concerts, but the Governor did not entirely remove the mandate that North Carolina was “Safer at Home” (Executive Order 141).

In an effort to provide transportation for essential personnel, the Mayor of Charlotte allowed the local public transit system (Charlotte Area Transit System; “CATS”) to continue operating, under a modified schedule and new safety precautions [11]. To encourage social distancing while still providing service to the public, CATS began blocking off seats that were less than the CDC’s recommended 6 feet apart on all of their vehicles [12]. From March 25, 2020, to June 8, 2020, the public could ride the light rail around the city free of charge [11]. On October 2, 2020, Governor Roy Cooper placed the state of NC under Phase III via Executive Order 169. This phase relaxed some heavier Phase II restrictions [13]. Between these restriction phases, extra safety measures were implemented on some of the higher-capacity public transportation vehicles. In addition to socially distant seating options [14], plexiglass barriers were installed on some vehicles between passenger seating sections. As an additional precaution, the Governor of North Carolina also mandated in Executive Order 180 that masks be worn at all times while riding any public transit vehicle [15]. At this time, North Carolina continues to operate in Phase III of restrictions.

For this study, we collected samples from high-touch surfaces at three different time points during the pandemic [Figure 1]. As the University was closing down and students were moving off campus, we swabbed high-touch surfaces on campus buses as well as surfaces in public areas where students frequently gather. In Phase II, during late July, we swabbed high-touch surfaces on CATS buses, Lynx light rail train cars, and paratransit vehicles. Samples were collected at the end of daily operations, before vehicles were cleaned and sanitized. In Phase III, during mid-November, we again collected samples from CATS vehicles, this time both before and after cleaning. Samples were analyzed using PCR-based molecular detection with standard CDC research use only (RUO) reagents [16]. While a small number of SARS-CoV-2 positive samples were detected among various sites during Phase I and Phase III, the vast majority of sites sampled were free from detectable contamination with SARS-CoV-2.

**Figure 1.**
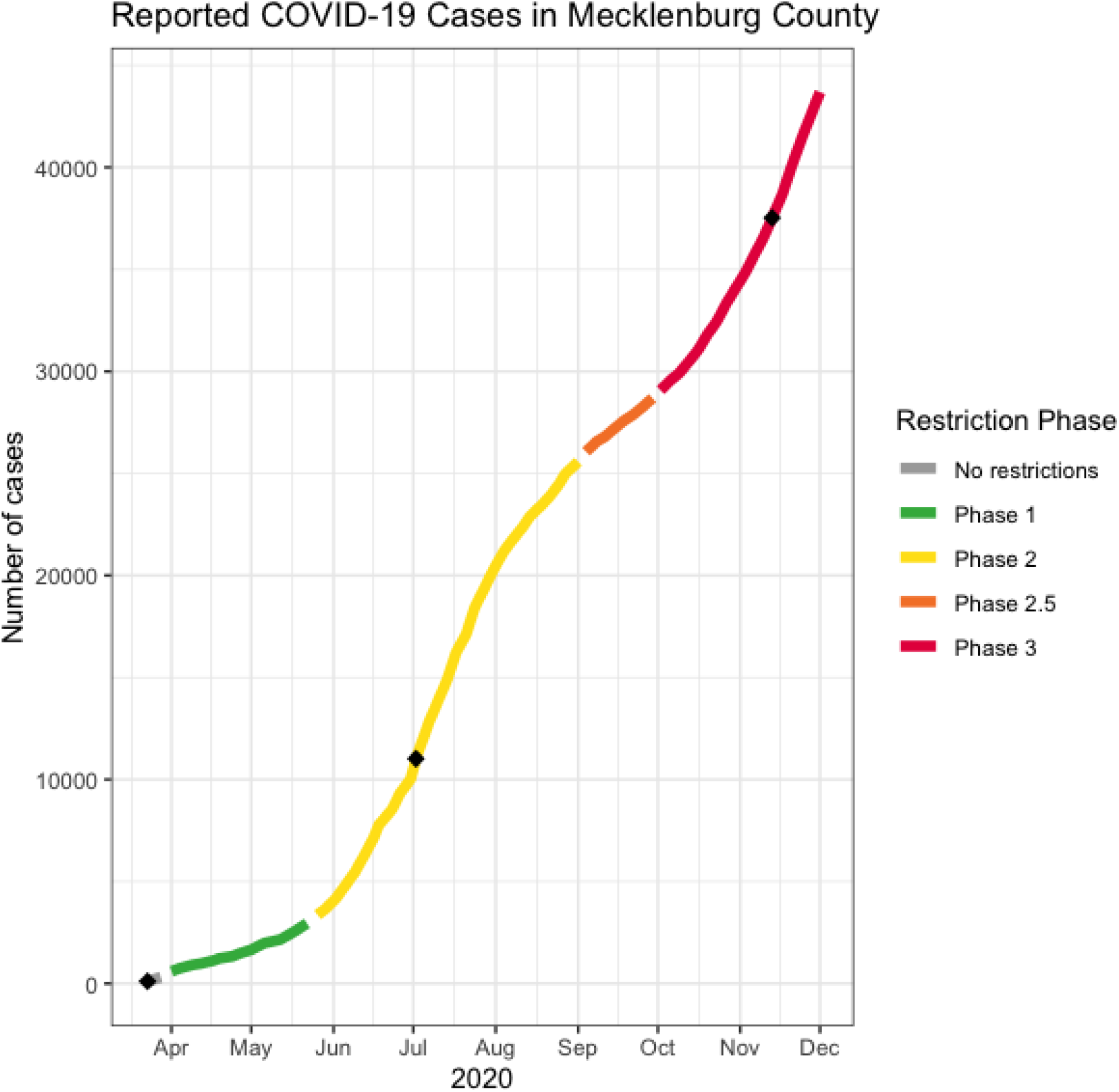
Graphical representation of Mecklenburg County’s reported COVID-19 cases over the 2020 pandemic period. The trendline of cases is segmented by the advent of restriction phases, and black diamond marks indicate when surface samples were collected.

## Materials and Methods

### Sample Collection and RNA Extraction

Sterile, PBS-soaked (Gibco, Waltham, MA) foam-tipped (Puritan Medical Products, Guilford, ME) swabs were used to collect samples from high-touch areas early in the pandemic on campus at the University of North Carolina at Charlotte (UNCC). Later, this swabbing protocol was repeated using polyester-tipped swabs (Texwipe, Kernersville, NC) on public transit vehicles before and after the completion of routine end-of-route sanitization procedures. As reagents became more readily available during the later sampling time points, each sample was replicated on three different public transit vehicles [Supplementary Table 1]. In all cases, immediately following the sample collection, the swabs were transferred into 15 mL Falcon tubes (Thermo Fisher Scientific, Waltham, MA) containing 1 mL of TRIzol™ (Invitrogen, Carlsbad, CA) and stored at 4°C until they could be processed. RNA was extracted and converted to cDNA using Applied Biosystems™ High Capacity cDNA Reverse Transcriptase MultiScribe kit (Thermo Fisher Scientific, Waltham, MA) in batches including 10 samples, a positive control (dilution of synthetic SARS-CoV-2 RNA 3 (LC528232.1), Twist Bioscience, San Francisco, CA), and a negative control (nuclease-free water).

### Collection Personnel Monitoring

Prior to collection of any samples, all collection personnel were asked to assess their health and trace contacts to the best of their abilities. Anyone that was feeling ill or had come in close contact with an infected individual was asked to refrain from sample collection and self-quarantine. To ensure that none of the able collection personnel were unintentionally contaminating the surface samples, each team member provided a saliva sample that was subject to the same analysis that all surface samples received.

### Detection of SARS-CoV-2 Using Endpoint PCR

In the early months of the pandemic, some of the necessary reagents for SARS-CoV-2 detection and quantification were reserved for healthcare research only, and we were initially limited to using endpoint PCR as a detection method. After the conversion of RNA to cDNA, we screened for the presence of SARS-CoV-2 RNA using Bioline’s MyFi™ kit (Meridian Bioscience, Cincinnati, OH) and IDT’s Research Use Only (RUO) N1 and N2 nucleocapsid gene primers. For each batch of 10 samples, a positive control (dilution of a plasmid containing the 2019-nCoV nucleocapsid gene; Integrated DNA Technologies, Coralville, IA) and negative control (nuclease-free water) was added. Samples were placed into a Bio-Rad T-100 thermocycler (Bio-Rad, Hercules, CA) and amplified using the following thermocycling conditions: 5 minutes at 98°C for initial denaturation, and 35 cycles of 5 seconds at 95°C for denaturation, 15 seconds at 55°C for annealing, and 30 seconds at 72°C for extension. Following PCR, 3μL of each amplified sample was then loaded into a pre-prepared 12% acrylamide gel. Samples that were positive for the presence of viral RNA were expected to show product for N1 and N2 at 72 and 67 base pairs, respectively.

### Detection of SARS-CoV-2 Using RT-qPCR

As quantitative PCR reagents became available for academic research again, we repeated analysis of early Phase I and Phase II samples using a quantitative PCR protocol. Once RNA was extracted from each sample, it was then converted to cDNA using the LunaScript™ Reverse Transcriptase SuperMix Kit (New England BioLabs, Inc., Ipswich, MA). Samples were analyzed in batches along with a positive control (dilution of synthetic SARS-CoV-2 RNA 1 (MT007544.1), Twist Bioscience, San Francisco, CA), and a negative control (nuclease-free water).

Each converted cDNA sample was placed in a sequencing plate in replicates of three, and received a master mix containing iTaq Universal Probes Reaction Mix (Bio-Rad, Hercules, CA), and IDT’s Research Use Only N1 or N2 nucleocapsid gene primers/fluorescent probes. Additional positive control (dilution of the plasmid containing 2019-nCoV nucleocapsid gene; Integrated DNA Technologies, Coralville, IA) and negative control (nuclease-free water) triplicates were added to the plate. The sequencing plate was sealed, placed into the CFX96 Touch Deep Well Real-Time PCR System (Bio-Rad, Hercules, CA) and then amplified using the following thermocycling conditions: 2 minutes at 25° for initiation, 2 minutes at 95° for polymerase activation, 45 cycles of 10 seconds at 95° for denaturation, and 30 seconds at 60° for extension.

### Spiked Virus Recovery Tests

In order to determine whether viral material applied to a surface could be recovered using the above swabbing approach and qPCR protocol, we executed a series of control experiments. 20 μL of a 1:5 dilution of both a 2019-nCoV nucleocapsid gene positive control (Integrated DNA Technologies, Coralville, IA) and a synthetic SARS-CoV-2 RNA (SARS-CoV-2 RNA 3 (LC528232.1), Twist Bioscience, San Francisco, CA) was applied to a 2×2 inch square glass plate inside of a biosafety hood. Samples were left to dry under the hood and after 30 minutes, were swabbed using both foam-tipped (Puritan Medical Products, Guilford, ME) and polyester-tipped (Texwipe, Kernersville, NC) swabs. The swabs were subsequently processed using the intended experimental protocol (detailed in *Sample Collection and RNA Extraction*). Following processing, the samples underwent conversion from RNA to cDNA and then RT-qPCR (detailed in *Detection of SARS-CoV-2 Using RT-qPCR* section) to determine their efficacy in recovering viral material from surfaces.

## Results

### Spiked Virus Recovery Tests

For the spiked viral recovery tests, initial concentrations of both IDT and Twist’s synthetic RNA ranged from 20-20000 copies/μL, respectively. We found that overall, we achieved more consistent recovery of material from glass plates spotted with the RNA-based SARS-CoV-2 positive control (Twist Bioscience) than with a plasmid based positive control (IDT) [Table 1]. Repeating the same set of experiments using the polyester-tipped swabs that became available to us in the later stages of the pandemic also showed positive recovery results. Of the surfaces spotted with the Twist artificial RNA, the polyester-tipped swabs recovered fractions ranging from 16-60% of viral RNA deposited, while recovery fractions using foam-tipped swabs were lower [Table 1]. A similar trend was observed whether N1 or N2 primers were used in the assay, but with a comparatively lower recovery rate across all swab types when the N2 primers were used [Table 1]. The lower recovery rate with the IDT positive control in the early endpoint experiments was likely due in part to the use of 35 cycles of amplification rather than 45 for early proof-of-concept tests. However, we did not pursue this issue further once the RNA-based control became available.

**Table 1.** The results of the spiked viral recovery tests, using both IDT and Twist’s synthetic RNA using IDT’s Research Use Only N1 and N2 nucleocapsid gene primers/fluorescent probes, respectively. All of the concentrations from the dilution series were tested; however, the table shows only those that gave a signal in the qPCR assay.

| N1 |  |  |  |  |  |  |
| --- | --- | --- | --- | --- | --- | --- |
| Swab Type | Initial Concentration (Copies/μL) | Initial Spiked Copy Number | Copies/Rxn | Copies/M<sup>2</sup> | Total Copies Recovered | Percentage Recovered (%) |
| Twist Foam Swab | 20000 | 400,000 | 1266.03 | 9.81x10<sup>6</sup> | 25320.69 | 6.33 |
| Twist Foam Swab | 2000 | 40,000 | 150.27 | 1.16x10<sup>6</sup> | 3005.41 | 7.51 |
| Twist Polyester Swab | 20000 | 400,000 | 3322.37 | 2.57x10<sup>7</sup> | 66447.31 | 16.61 |
| IDT Polyester Swab | 4000 | 80,000 | 387.70 | 3.00x10<sup>6</sup> | 7754.03 | 9.70 |
| Twist Polyester Swab | 2000 | 40,000 | 1200.33 | 9.30x10<sup>6</sup> | 24006.62 | 60.02 |
| Twist Polyester Swab | 200 | 4,000 | 78.56 | 6.08x10<sup>5</sup> | 1571.23 | 39.28 |
| N2 |  |  |  |  |  |  |
| Swab Type | Initial Concentration (Copies/μL) | Initial Spiked Copy Number | Copies/Rxn | Copies/M<sup>2</sup> | Total Copies Recovered | Percentage Recovered (%) |
| Twist Foam Swab | 20,000 | 400,000 | 987.63 | 3.83x10<sup>5</sup> | 19752.52 | 4.94 |
| Twist Foam Swab | 2,000 | 40,000 | 44.33 | 1.72x10<sup>4</sup> | 886.51 | 2.22 |
| Twist Polyester Swab | 20,000 | 400,000 | 1583.18 | 6.13x10<sup>5</sup> | 31663.66 | 7.92 |
| IDT Polyester Swab | 4,000 | 80,000 | 92.52 | 3.59x10<sup>4</sup> | 1850.37 | 2.31 |
| Twist Polyester Swab | 2,000 | 40,000 | 455.21 | 1.76x10<sup>5</sup> | 9104.18 | 22.76 |

Using both the qPCR data and the spiked viral RNA recovery tests, we were able to calculate the number of viral RNA copies present per swab, per reaction, and per swabbed surface [Table 2]. A recent virological assessment of SARS-CoV-2 positive patients’ coughed/sneezed sputum found an average viral load of 7 × 10^6^ copies/mL, with a maximum of 2.35 × 10^6^ copies/mL [17]. The amount of Twist or IDT control that we used in the spiked recovery test was equivalent to a range between 2 × 10^4^ copies/mL to 4 × 10^8^ copies/mL, making it a representative proxy for SARS-CoV-2 positive sputum in terms of the number of viral copies deposited in fluid form by a cough or sneeze. We measured RNA recovered from areas that were likely to contain evaporated, settled sputum droplets, and our measurements were calculated in units of area. The site with the largest number of recovered viral RNA copies/M^2^ when using the N1 primer/probe set was 1.6 x 105; a set of stainless steel grab poles on the right and left of the entryway door on the second sampled train. When using the N2 primer/probe set, this same site showed a viral RNA copy recovery of 1.7 x 104 copies/M^2^. The remaining two sites showed less variation in copies per area, with the vinyl grab ring showing 4.7 x 104 copies/M^2^ using the N1 primer/probe set, and the vinyl-covered stop request line showing 2.3 x 104 copies/M^2^ using the N2 primer/probe set.

**Table 2.** SARS-CoV-2 viral copies recovered, calculated from N1 and N2 positive public transit samples collected in Phase III of sampling, respectively. From the mean Cq value, viral copies per reaction, total number of viral copies recovered from the experimental swab, and detectable viral copies per meter squared were computed.

| N1 |  |  |  |  |
| --- | --- | --- | --- | --- |
| Site | Mean Cq | Copies/rxn | Total Copies Recovered from Swab | Copies/M<sup>2</sup> |
| 1T2.2 | 35.11 | 20.75 | 415.00 | 1.61x10<sup>5</sup> |
| GR2 | 36.82 | 6.18 | 123.52 | 4.78x10<sup>4</sup> |
| N2 |  |  |  |  |
| Site | Mean Cq | Copies/rxn | Total Copies Recovered from Swab | Copies/M<sup>2</sup> |
| 1T2.2 | 38.15 | 2.25 | 45.07 | 1.75x10<sup>4</sup> |
| 2B5.1 | 37.74 | 3.02 | 60.47 | 2.34x10<sup>4</sup> |

### Early Pandemic Campus Samples

At the beginning of the pandemic, we were able to demonstrate that the endpoint PCR method performed as expected [Supplementary Figure 1], and to successfully detect the presence of SARS-CoV-2 RNA [Figure 2] from surface samples collected on campus using endpoint PCR alone. This method detected SARS-CoV-2 in 11 of 70 samples using the RUO N2 primers, from locations on campus at UNCC [Table 3; Table 4; Figure 2]. Of the 11 that were deemed positive using the endpoint PCR detection method, 3 of those were later confirmed positive for SARS-CoV-2 using the qPCR detection method [Table 5].

**Table 3.** 14 samples were obtained from the campus bus transportation system in the first phase of surface sampling. Of these 14 samples, two tested positive for the presence of SARS-CoV-2 using IDT’s N2 primer/probe set in an endpoint PCR analysis.

| Campus Bus Samples |  |  |
| --- | --- | --- |
| Sample Name | Collection Date | Sample +/- |
| **Bus 1; Left side of door; handle** | **3/25/20** | **Positive** |
| Bus 1; Right side of door; handle | 3/25/20 | Negative |
| Bus 1; Horizontal railing | 3/25/20 | Negative |
| Bus 1; Plastic grab rings | 3/25/20 | Negative |
| Bus 1; Cloth seats | 3/25/20 | Negative |
| Bus 2; Left side of door; handle | 3/25/20 | Negative |
| Bus 2; Right side of door; handle | 3/25/20 | Negative |
| **Bus 2; Horizontal railing** | **3/25/20** | **Positive** |
| Bus 3; Right side of door; handle | 3/27/20 | Negative |
| Bus 3; Left side of door; handle | 3/27/20 | Negative |
| Bus 3; Stop request lines; front/middle | 3/27/20 | Negative |
| Bus 3; Stop request lines; back | 3/27/20 | Negative |
| Bus 3; Cloth seats | 3/27/20 | Negative |
| Bus 3; Plastic grab rings | 3/27/20 | Negative |

**Table 4.** 56 samples were obtained from various sites on campus at UNCC in the first phase of surface sampling. Of these 56 samples, 11 samples tested positive for the presence of SARS-CoV-2 using IDT’s N2 primer/probe set in an endpoint PCR analysis.

| Campus Samples |  |  |
| --- | --- | --- |
| Sample Name | Collection Date | Sample +/- |
| Residence Hall 1; Left Hand door handle | 3/23/20 | Negative |
| Residence Hall 1; Right Hand door handle | 3/23/20 | Negative |
| Residence Hall 2; Left Hand door handle | 3/23/20 | Negative |
| Residence Hall 2; Right Hand door handle | 3/23/20 | Negative |
| Residence Hall 3; Main door handles | 3/23/20 | Negative |
| Residence Hall 4; Upstairs corner door handles | 3/23/20 | Negative |
| Residence Hall 4; Downstairs corner door handles | 3/23/20 | Negative |
| **Residence Hall 5; Upstairs corner door handles** | **3/23/20** | **Positive** |
| Residence Hall 5; Downstairs corner door handles | 3/23/20 | Negative |
| Student Union; Exterior; Right Hand doors 1 | 3/23/20 | Negative |
| Student Union; Exterior; Right Hand doors 2 | 3/23/20 | Negative |
| Student Union; Interior; Right Hand doors 1 | 3/23/20 | Negative |
| Student Union; Interior; Right Hand doors 2 | 3/23/20 | Negative |
| Student Union; Exterior; Left Hand doors 3 | 3/23/20 | Negative |
| Student Union; Interior; Left Hand doors 3 | 3/23/20 | Negative |
| Student Union; ATM | 3/23/20 | Negative |
| Student Union; Convenience store; Refrigerators | 3/23/20 | Negative |
| **Student Union; Lounge** | **3/23/20** | **Positive** |
| Library; Exterior doors 1 | 3/23/20 | Negative |
| Library; Interior doors 1 | 3/23/20 | Negative |
| Library; Detector exit 1 | 3/23/20 | Negative |
| Library; Exterior doors 2 | 3/23/20 | Negative |
| Library; Exterior doors 3 | 3/23/20 | Negative |
| Library; Interior doors 2 | 3/23/20 | Negative |
| **Library; Interior doors 3** | **3/23/20** | **Positive** |
| ATM 1 | 3/23/20 | Negative |
| ATM 2 | 3/23/20 | Negative |
| ATM 3 | 3/23/20 | Negative |
| Student Health Center; Lower entrance doors | 3/23/20 | Negative |
| **Student Health Center; Main entrance doors** | **3/23/20** | **Positive** |
| Check-in computer 1 | 3/23/20 | Negative |
| Check-in computer 2 | 3/23/20 | Negative |
| Check-in computer 3 | 3/23/20 | Negative |
| Check-in computer 5 | 3/23/20 | Negative |
| Residence Hall 1; Left Hand door handle | 3/25/20 | Negative |
| **Residence Hall 1; Right Hand door handle** | **3/25/20** | **Positive** |
| Residence Hall 1; Main door | 3/25/20 | Negative |
| **Residence Hall 2; Main door** | **3/25/20** | **Positive** |
| **Residence Hall 2; Left Hand door handle** | **3/25/20** | **Positive** |
| Residence Hall 2; Right Hand door handle | 3/25/20 | Negative |
| Residence Hall 3; Main door handles | 3/25/20 | Negative |
| Residence Hall 4; Upstairs corner door handles | 3/25/20 | Negative |
| **Residence Hall 4; Downstairs corner door handles** | **3/25/20** | **Positive** |
| **Residence Hall 5; Upstairs corner door handles** | **3/25/20** | **Positive** |
| **Residence Hall 5; Downstairs corner door handles** | **3/25/20** | **Positive** |
| Student Union; Exterior; Right Hand doors 1 | 3/25/20 | Negative |
| Student Union; Exterior; Right Hand doors 2 | 3/25/20 | Negative |
| **Student Union; Interior; Right Hand doors 1** | **3/25/20** | **Positive** |
| Student Union; Interior; Right Hand doors 2 | 3/25/20 | Negative |
| Student Union; Exterior; Left Hand doors 3 | 3/25/20 | Negative |
| Student Union; Interior; Left Hand doors 3 | 3/25/20 | Negative |
| **Student Union; ATM** | **3/25/20** | **Positive** |
| Student Union; Convenience store; Refrigerators | 3/25/20 | Negative |
| **Student Union; Acrylic-sealed pool cues** | **3/25/20** | **Positive** |
| Library; Exterior doors 1 | 3/25/20 | Negative |
| Library; Interior doors 1 | 3/25/20 | Negative |

**Table 5.** Of collected bus and campus sites that tested positive using the endpoint PCR method with IDT’s N2 primer/probe set, 3 sites tested positive for the presence of SARS-CoV-2 using IDT’s N2 primer/probe set with qPCR analysis (at least two of three wells reporting positive cycle threshold (Cq) values are considered positive).

| **Sample** | **Date Collected** | **Cq Mean** | **Cq Std. Dev.** | **Sample +/-** |
| --- | --- | --- | --- | --- |
| Residence Hall 5; Upstairs corner door handles | 3/23/20 | 0 | 0 | Negative |
| Bus 2; Horizontal railing | 3/25/20 | 0 | 0 | Negative |
| Residence Hall 5; Downstairs corner door handles | 3/23/20 | 0 | 0 | Negative |
| Student Union; Lounge | 3/23/20 | 0 | 0 | Negative |
| Residence Hall 1; Right Hand door handle | 3/25/20 | 0 | 0 | Negative |
| Student Union; Interior; Right Hand doors 1 | 3/25/20 | 0 | 0 | Negative |
| Library; Interior doors 3 | 3/23/20 | 0 | 0 | Negative |
| Residence Hall 2; Main door | 3/25/20 | 0 | 0 | Negative |
| Student Union, ATM | 3/25/20 | 0 | 0 | Negative |
| **Student Health Center; Main entrance doors** | **3/23/20** | **34.05** | **0.306** | **Positive** |
| **Residence Hall 2; Left Hand door handle** | **3/25/20** | **32.98** | **0.489** | **Positive** |
| Student Union; Acrylic-sealed pool sticks | 3/25/20 | 0 | 0 | Negative |
| Bus 1; Left side of door; handle | 3/25/20 | 0 | 0 | Negative |
| **Residence Hall 4; Downstairs corner door handles** | **3/25/20** | **34.78** | **0.11** | **Positive** |

**Figure 2.**
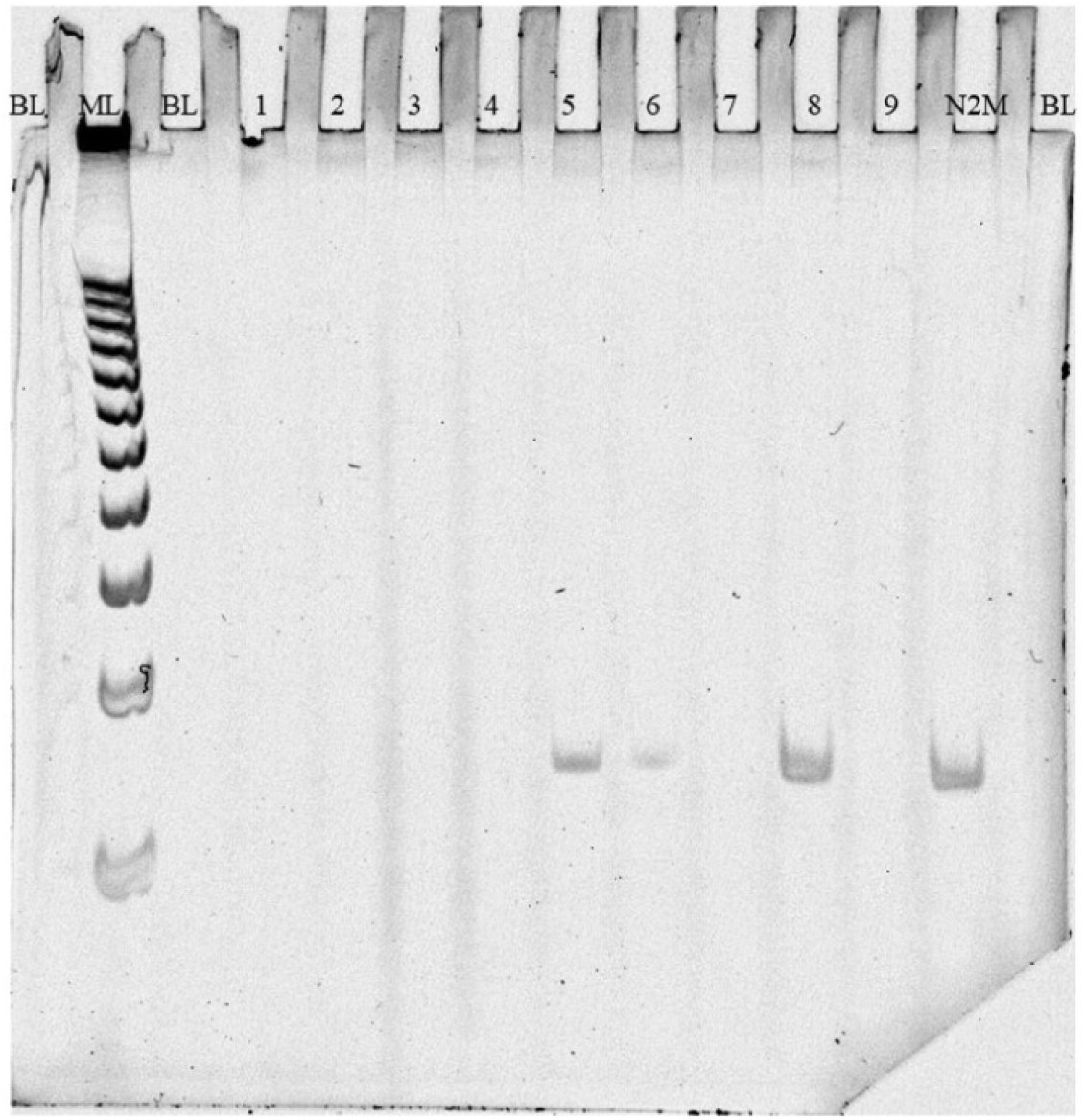
Example acrylamide gel image depicting all components indicative of a successful experiment. In this image, the first and last lanes labeled “BL” are negative controls loaded with nuclease-free water, and the lane labeled “ML” contains a 25 base pair ladder. IDT’s 2019-nCov’s specific N2 primer produces a PCR product that is 67 base pairs in length, seen in lane “N2M” for positive control purposes. Bands at the same location are considered to have tested positive for SARS-CoV-2, seen in lanes 5 and 6. These lanes contain the positive PCR product of experimental samples collected from stainless steel door handles to a student residence hall, as well as the university’s heavily populated Student Union.

### Phase II and III; Public Transit Samples

Of the 51 samples collected from CATS vehicles during the July sampling event in Phase II, none were positive for the presence of viral particles using both endpoint PCR analysis [Supplementary Figure 2] as well as qPCR with IDT’s N2 primer/probe set [Supplementary Table 3]. Of the 116 samples collected from CATS vehicles during the November sampling event in Phase III, 3 of the collected samples tested positive for the presence of SARS-CoV-2 RNA with the N1, N2, or both primers [Table 5].

In order for a sample to be considered positive, each of the three replicates from the qPCR assay must show that it has crossed the cycle threshold (Cq) at or below 40 cycles per CDC guidelines [15]. The samples collected on the stop request line on the second bus showed Cq values consistent with a positive in all three wells for the N2 primer pre-sanitization [Figure 3; Table 6], and the grab rings on the samples collected from the second train showed Cq values consistent with a positive for all three wells for the N1 primer post-sanitization [Figure 4; Table 6]. The sample collected on the grab poles on the right/left of the entryway doors on the first train showed Cq values consistent with a positive result in all three wells for both the N1 and N2 primer, post-sanitization [Figure 4; Table 6]. None of the samples collected from the paratransit vehicles showed any signal in the qPCR analyses using IDT’s N1 or N2 primer [Figure 5; Supplementary Table 2].

**Figure 3.**
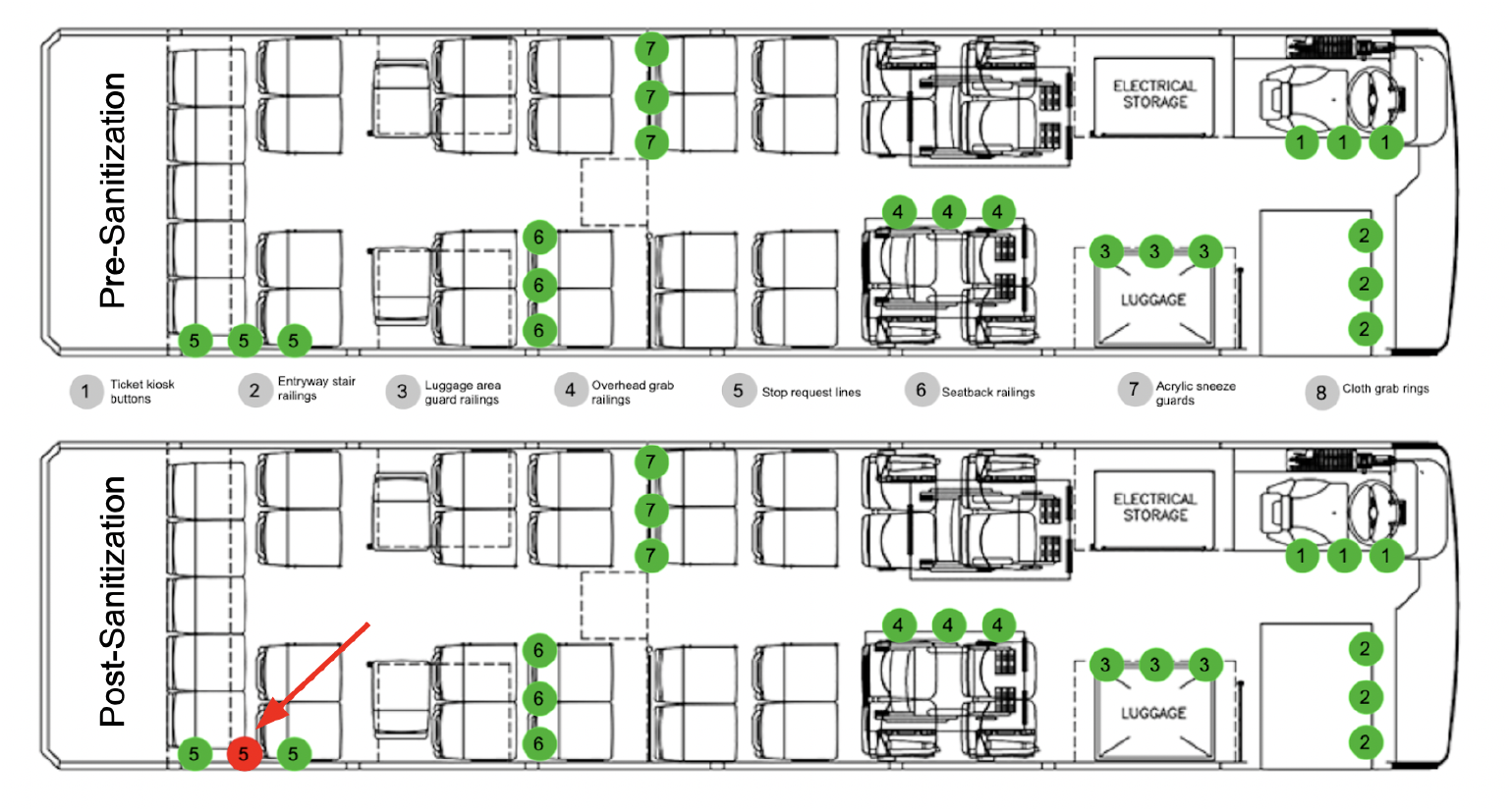
Illustration of sampling locations on the CATS buses [18]. Sites are labeled 1-8 on each vehicle, both before and after routine sanitization. A site with a green dot indicates that there was no presence of SARS-CoV-2 RNA recovered from the surface, and a site with a red dot indicates that there was presence of SARS-CoV-2 RNA recovered from the surface. Here, the stop request line at the back of bus 2 after cleaning tested positive for the presence of SARS-CoV-2 RNA.

**Table 6.**
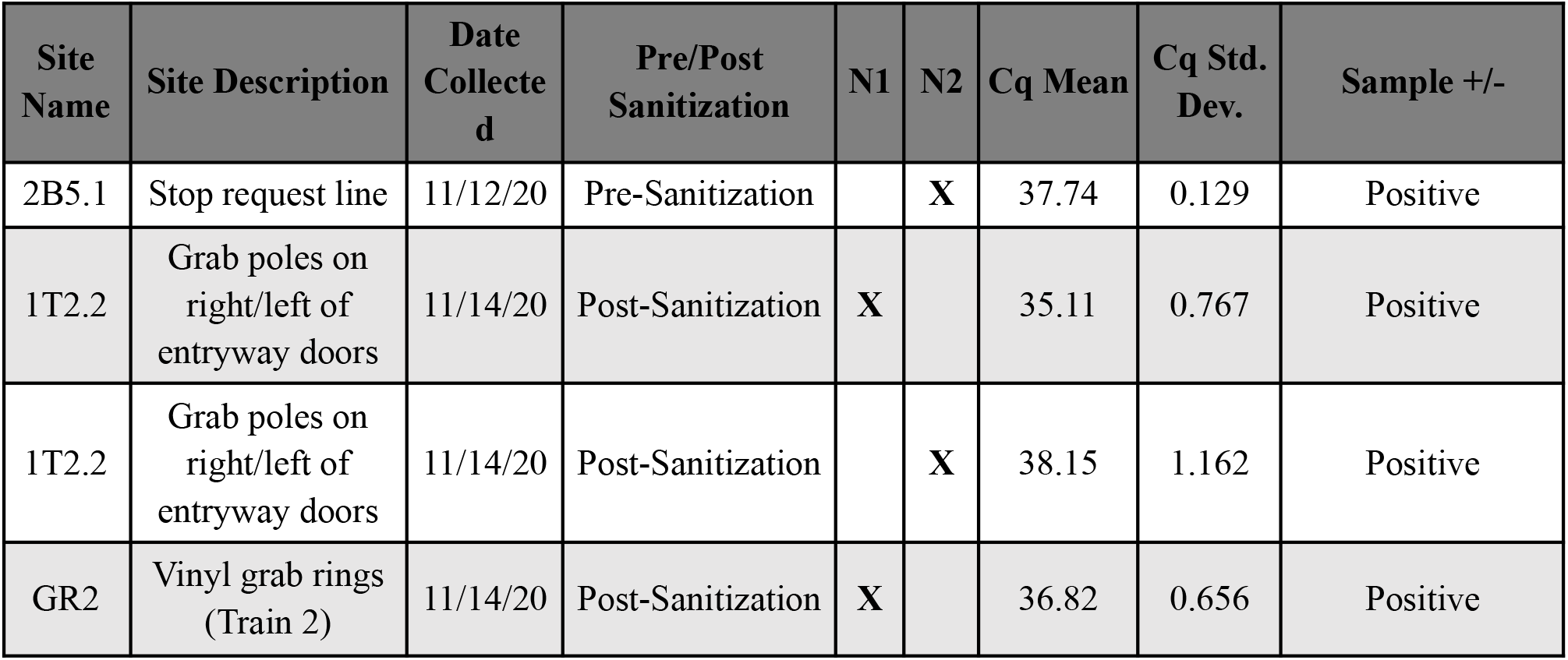
The three experimental samples indicating the presence of SARS-CoV-2 RNA in iteration 2 of the experiment, performed in Phase III.

| Site Name | Site Description | Date Collected | Pre/Post Sanitization | N1 | N2 | Cq Mean | Cq Std. Dev. | Sample +/- |
| --- | --- | --- | --- | --- | --- | --- | --- | --- |
| 2B5.1 | Stop request line | 11/12/20 | Pre-Sanitization |  | X | 37.74 | 0.129 | Positive |
| 1T2.2 | Grab poles on right/left of entryway doors | 11/14/20 | Post-Sanitization | X |  | 35.11 | 0.767 | Positive |
| 1T2.2 | Grab poles on right/left of entryway doors | 11/14/20 | Post-Sanitization |  | X | 38.15 | 1.162 | Positive |
| GR2 | Vinyl grab rings (Train 2) | 11/14/20 | Post-Sanitization | X |  | 36.82 | 0.656 | Positive |

**Figure 4.**
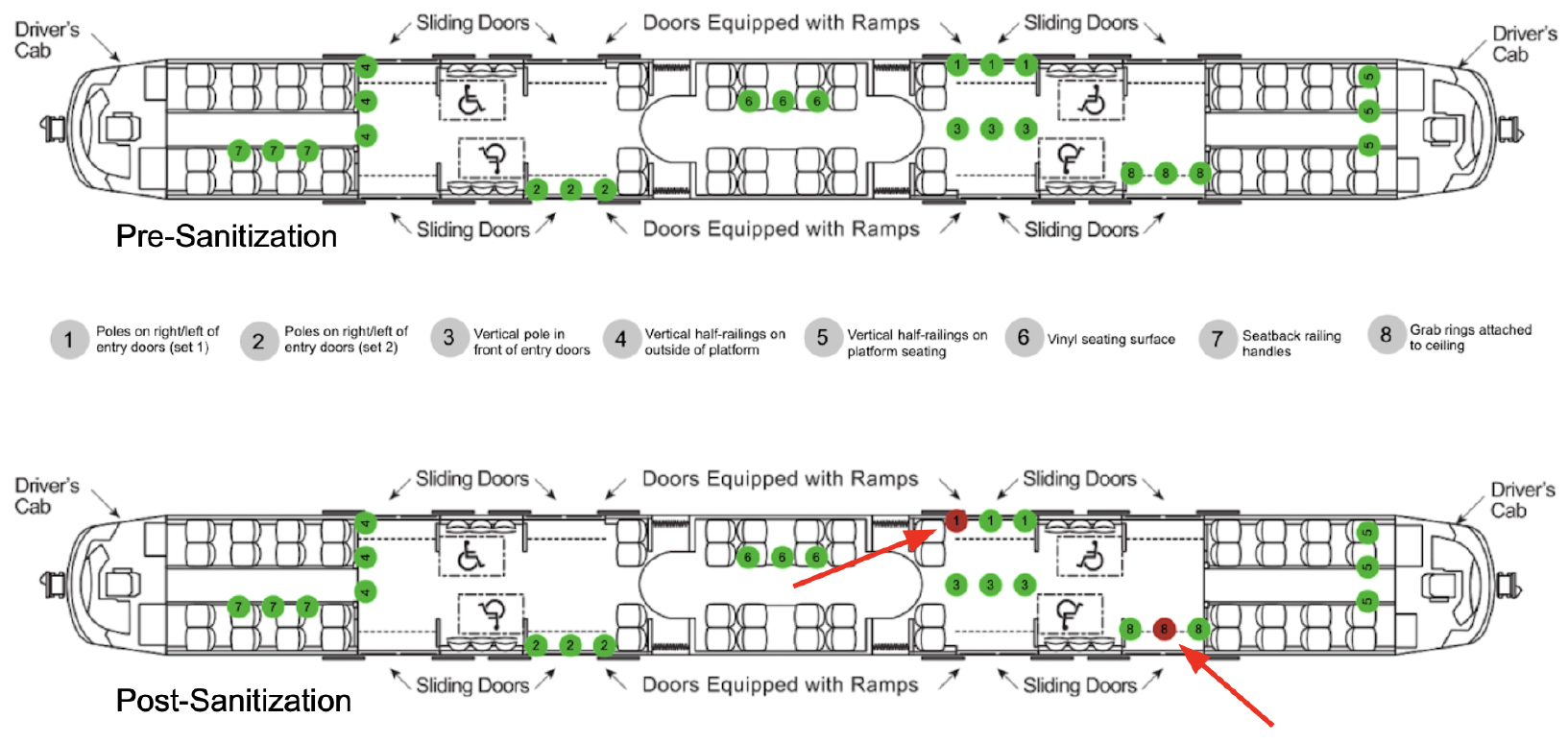
Illustration of sampling locations on the CATS Light Rail trains [19]. Sites are labeled 1-8 on each vehicle, both before and after routine sanitization. A site with a green dot indicates that there was no presence of SARS-CoV-2 RNA recovered from the surface, and a site with a red dot indicates that there was presence of SARS-CoV-2 RNA recovered from the surface. Here, the poles on the right/left of the entry doors on train 1 and the grab rings attached to the ceiling on train 2 tested positive for the presence of SARS-CoV-2 RNA.

**Figure 5.**
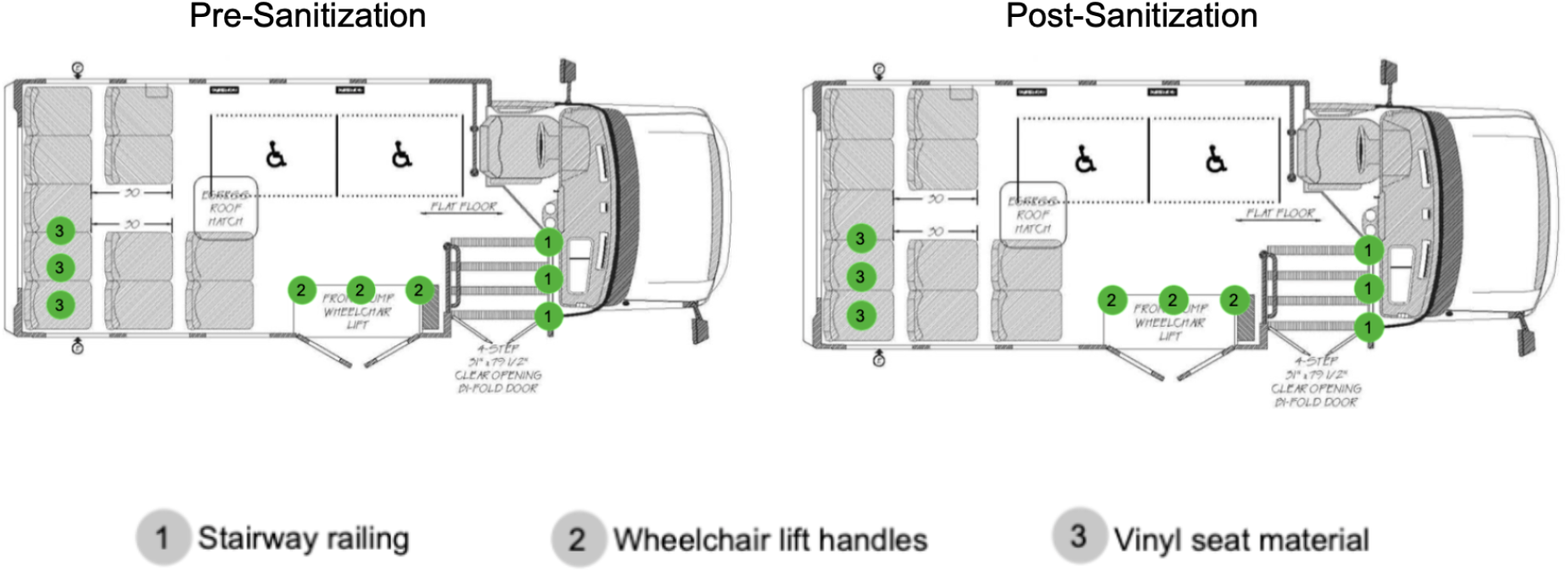
Illustration of sampling locations on the CATS paratransit vehicles [20]. Sites are labeled 1-3 on each vehicle, both before and after routine sanitization. A site with a green dot indicates that there was no presence of SARS-CoV-2 RNA recovered from the surface, and a site with a red dot indicates that there was presence of SARS-CoV-2 RNA recovered from the surface.

## Discussion

Beginning this experiment during a time when very little was known about SARS-CoV-2 gave our team a unique opportunity to use our research to inform the public. During the initial stages of the pandemic, decontaminating high-touch surfaces and practicing proper hand hygiene was one of the very first pieces of guidance that health agencies recommended as a defense against the spread of the virus [20], and this experiment was designed to assess that risk in public settings at a time when surface contamination had mainly been studied in high-risk hospital settings.

### Recovery tests validate the sampling approach

In spring 2020, swabs for sample collection were nearly impossible to find, constraining our choice of materials. We were later able to perform a variety of recovery tests using different swabs and materials to demonstrate that our sampling and molecular detection protocols were likely to be able to recover detectable virus in amounts deposited by a sneeze or a touch from a contaminated hand. Table 1 shows that the foam swabs that were used were able to recover viral RNA from surfaces using our swabbing protocol and Twist’s artificial SARS-CoV-2 RNA, albeit at low percentages of recovery.

### Both endpoint and quantitative PCR are capable of detecting surface contamination

Prior to analyzing surface samples with the endpoint PCR protocol, we tested batches of positive and negative controls with both the N1 and N2 primer sets. The N1 primer set yielded a number of false positives in endpoint PCR, while the N2 primer set did not. We therefore chose to use the N2 primer set in all remaining endpoint PCR tests. The 11 sites that tested positive in endpoint PCR were a variety of surface types, but a majority of them were stainless steel handles [Table 3]. Things like stainless steel door handles and grab rails are likely to be frequently touched by multiple individuals, with little opportunity for sanitization in between touches during busy times. We know now that SARS-CoV-2 RNA can remain on stainless steel surfaces for up to 72 hours after exposure [6], so finding viral material on these types of surfaces is not an unexpected outcome.

While not approved for use as a clinical diagnostic tool, the endpoint PCR detection protocol used in our early tests was capable of detecting viral material from surfaces. The early results convinced us that testing surfaces on regional public transit would be of interest. By the time we collected samples in Phase II, stringent cleaning procedures had been implemented by CATS, and PCR results were reassuringly negative at all sites. Follow-up qPCR tests of the same samples corroborated that finding, and the qPCR protocol used was also able to detect viral contamination when it was present in Phase III of the study. In the surface sample collection during Phase III, as in Phase II, SARS-CoV-2 RNA was not recovered from surfaces prior to cleaning. However, virus was detected in several sites after sanitization was completed. This suggested one of the following scenarios: these areas of high contact were not thoroughly sanitized at the end of the day (which our data did not support), or perhaps that a member of the cleaning crew could have been, unknowingly, spreading viral RNA during the sanitization process. After reporting our findings to CATS leadership, they confirmed that there had been an asymptomatic individual working in vehicle maintenance on the date samples were collected.

Despite the greater sensitivity of qPCR, however, Cq values values for surface samples hovered around the limit of detection of the assay (Cq value of 37.68; 1.5 x 104 copies/L), and therefore only three of the campus sites that tested positive in Phase I using endpoint PCR were unambiguously confirmed in qPCR using the SARS-CoV-2 N2 primer/probe set [Table 4]. Based on these outcomes, we believe it is likely that qPCR is not the ideal choice of method for these very low-concentration samples, as has been established in other applications, and that direct quantification of copies present using ddPCR, being a more sensitive endpoint method, would likely be a better choice for analysis of future surface samples [21]. Samples collected for Phase II and III of this experiment have also been contributed to the International MetaSUB Project’s MetaCov study [22], and results from sequencing of the collected material are likely to shed further light on the sensitivity and desirability of qPCR as an assay for surface detection of viral material.

### Detectable SARS-CoV-2 contamination on public high touch surfaces is infrequent

Overall, the occurrence of recovered RNA from surface samples was low in comparison to the total number of surfaces that were swabbed, especially in the later stages of the pandemic when aggressive cleaning protocols had been established. While we know that the primary mode of transmission of the virus is through airborne particles, our findings do indicate that detectable RNA can still occasionally be recovered from high-touch surfaces, especially if the contamination is recent. Precautions against touching face or eyes after contact with public surfaces remain reasonable, but the lack of detectable SARS-CoV-2 on public transit surfaces even prior to nightly cleaning suggests that current sanitation practices are adequate to minimize the chance of contact with viral material.

## Data Availability

All data is presented in supplementary files included in this submission.

## Supplementary Information

**Supplementary Table 1.**
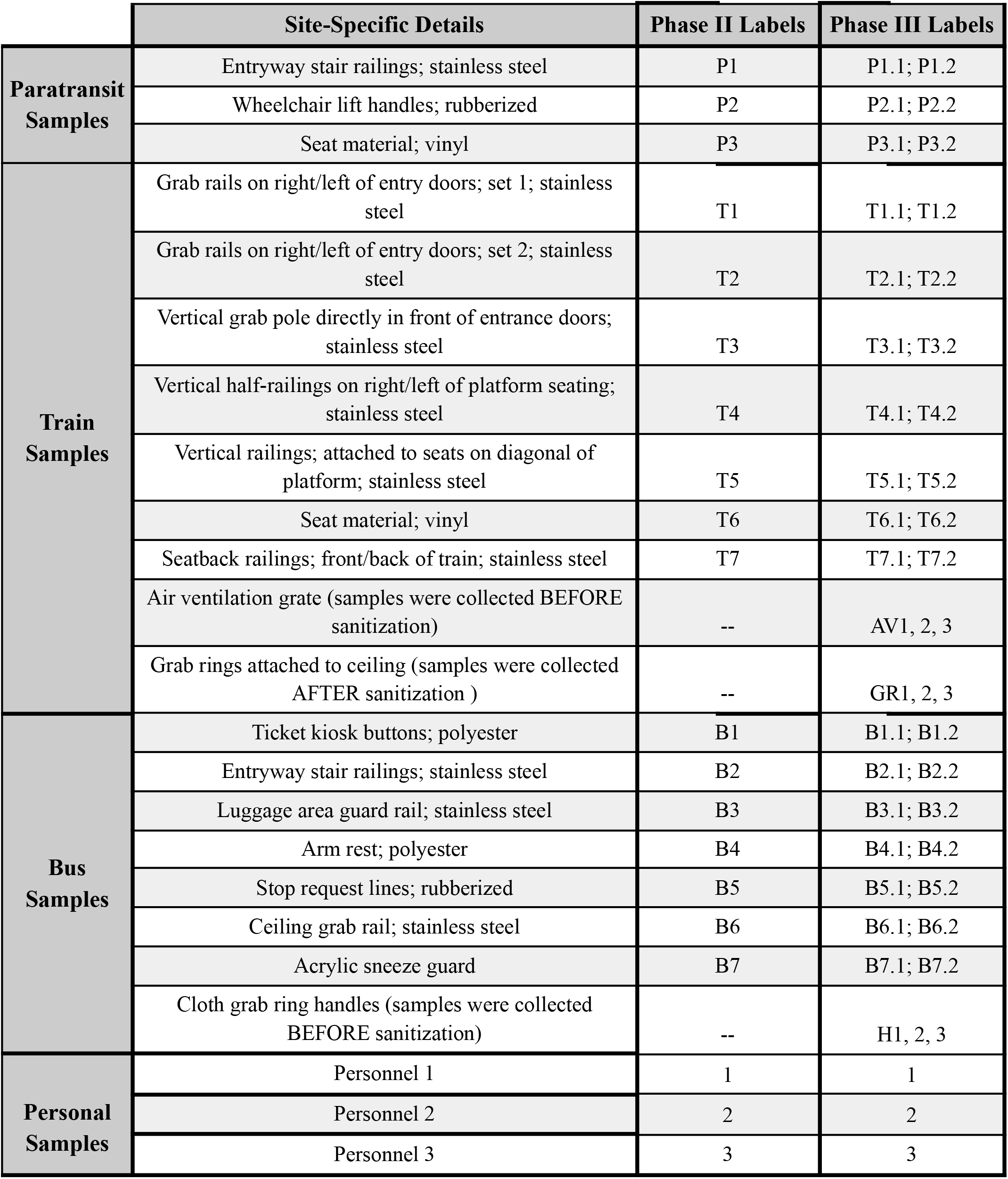
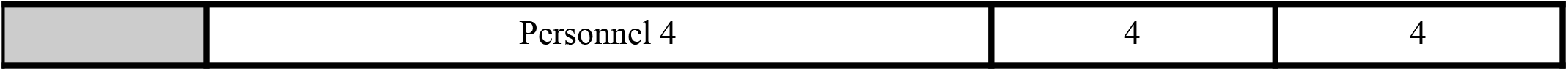
Sampling locations with site-specific metadata. Samples collected in Phase II were sampled prior to vehicle sanitization, and samples collected in Phase III were collected before (denoted by a .1) and after (denoted by a .2) vehicle sanitization.

**Supplementary Figure 1.**
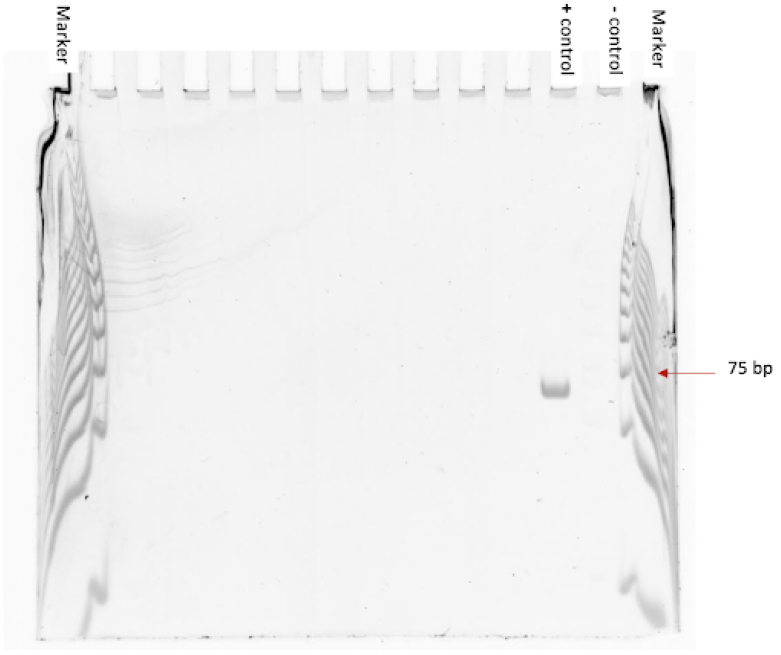
Example gel image depicting all components indicative of a successful experiment. In this image, the first and last lanes contain the 25 base pair ladder, the 75bp band is indicated. The 2019-nCoV’s specific N1 primer produces a PCR product that is 72bp in length, and the 2019-nCov’s specific N2 primer produces a PCR product that is 67bp in length. The second to last lane containing the negative control is completely blank, and the positive control shows a band around the 75 base pair mark. A band here indicates a positive result; in the case of our experiment, this would indicate that there is presence of SARS-CoV-2 viral RNA.

**Supplementary Figure 2.**
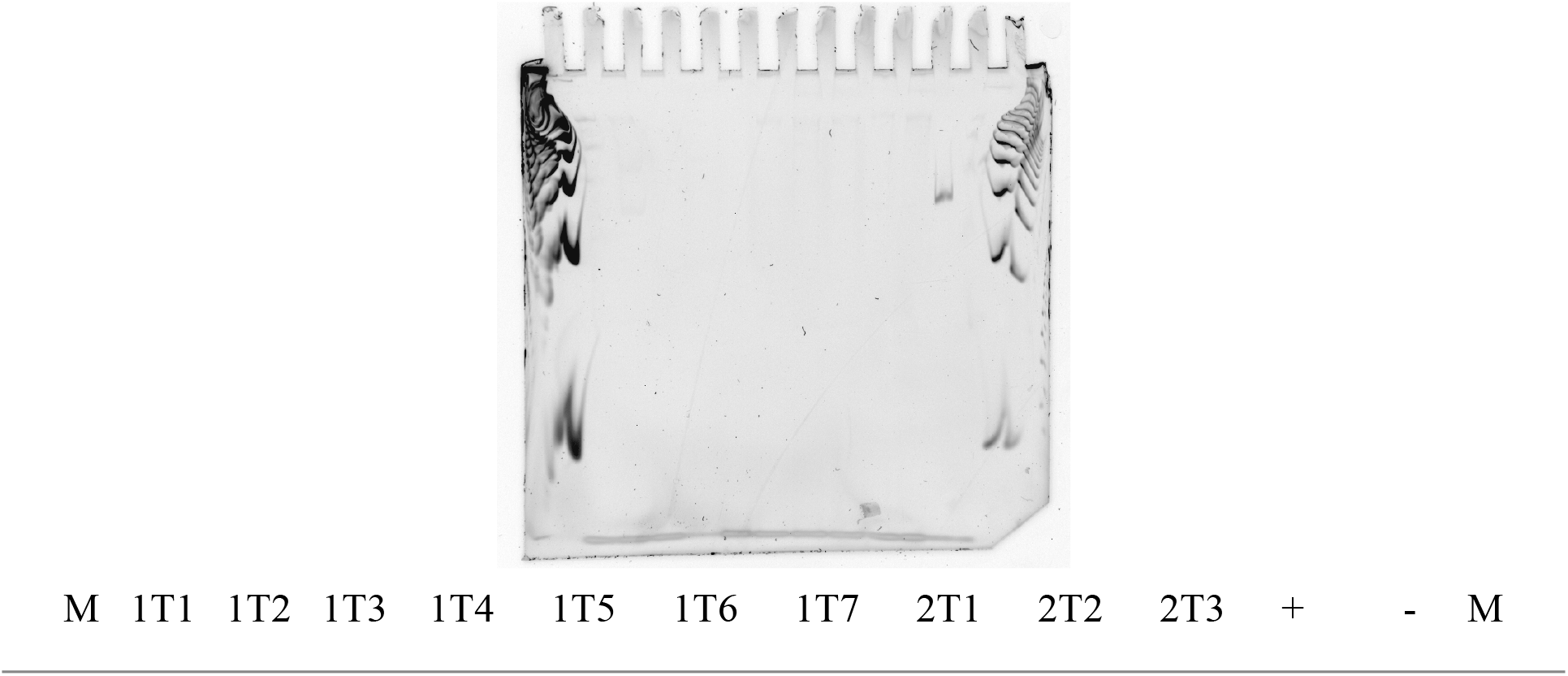
Plate 1; row A (all gels for public transit vehicles in Phase II look like this)

**Supplementary Table 2.**
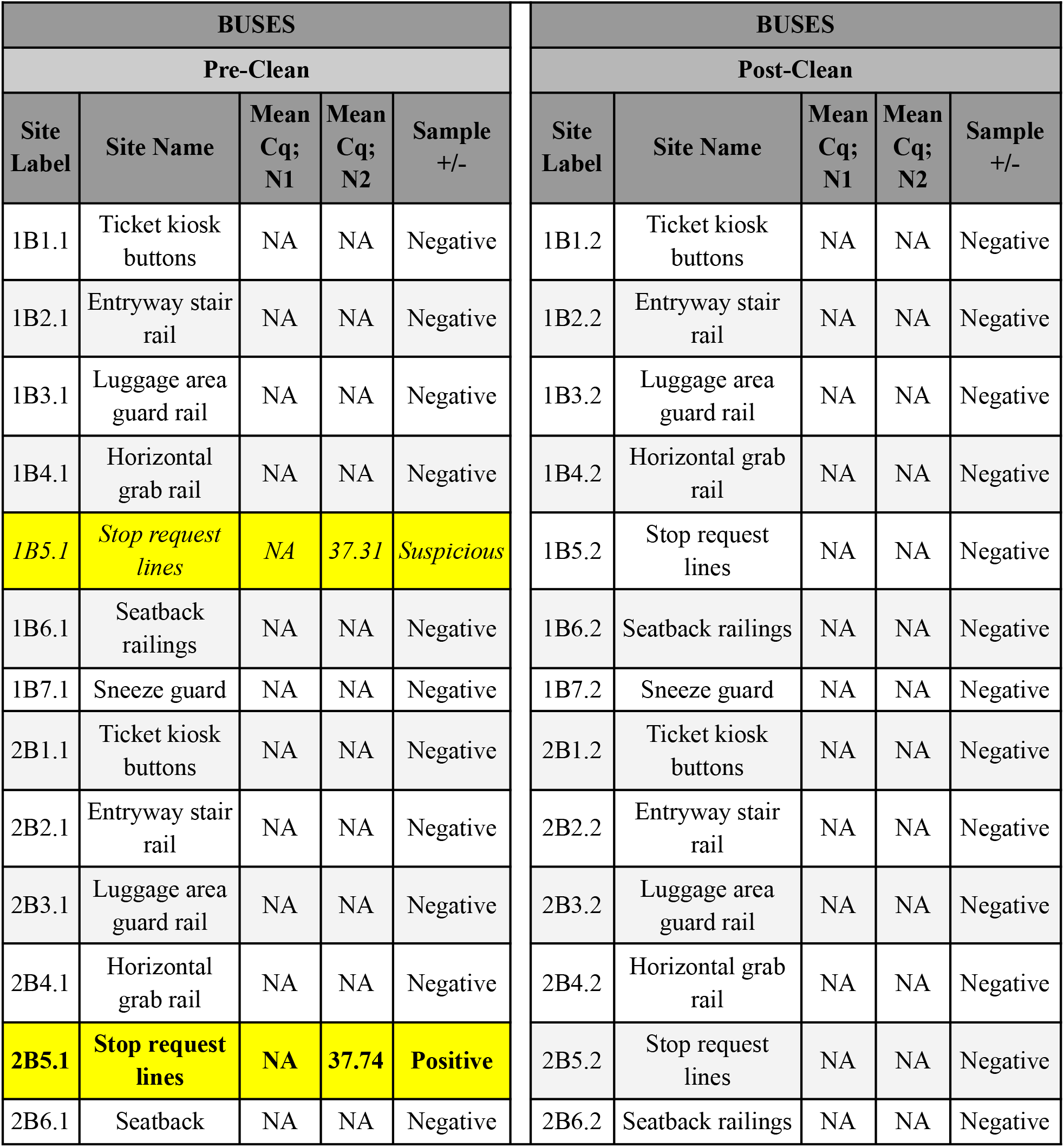
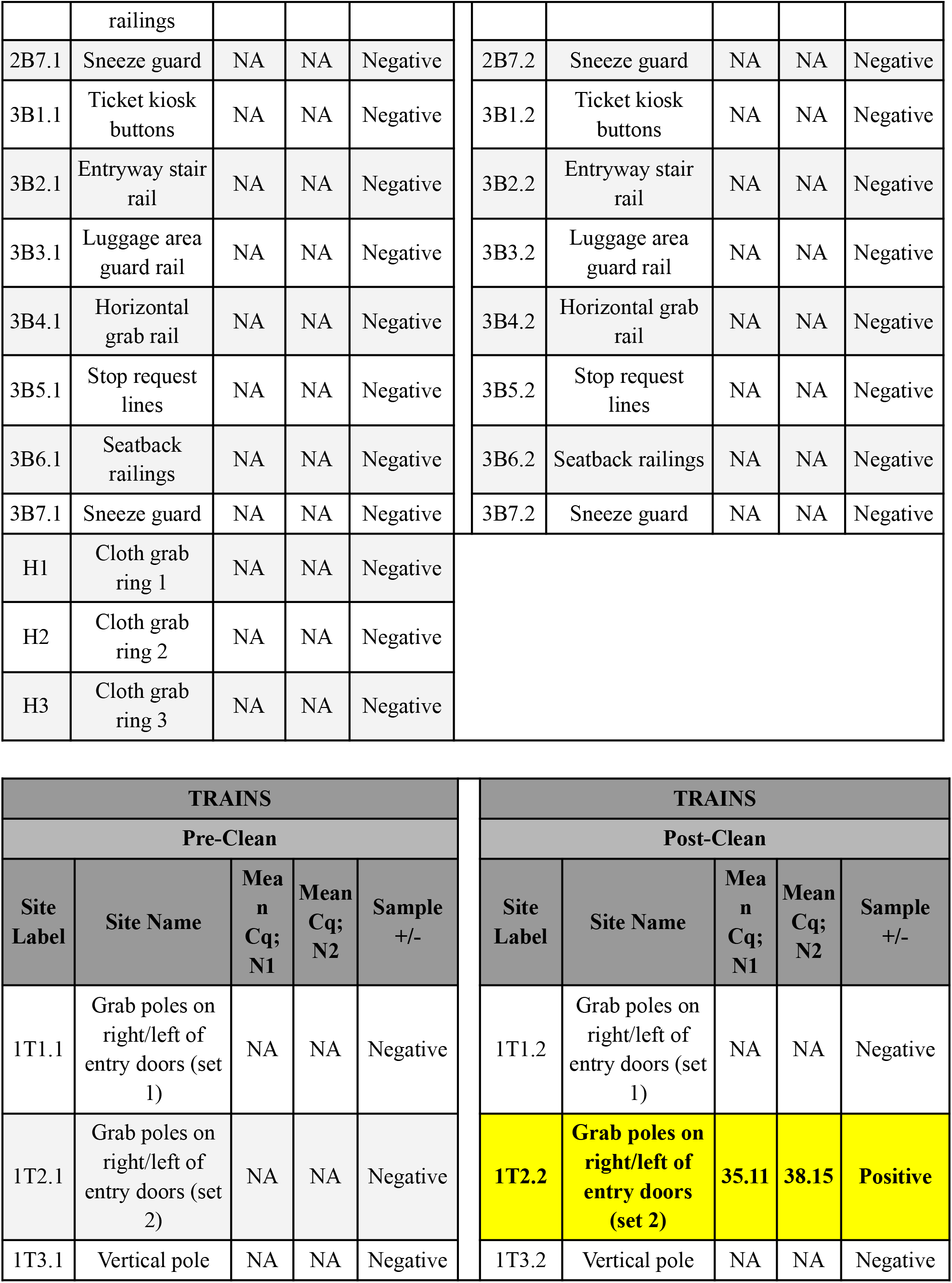
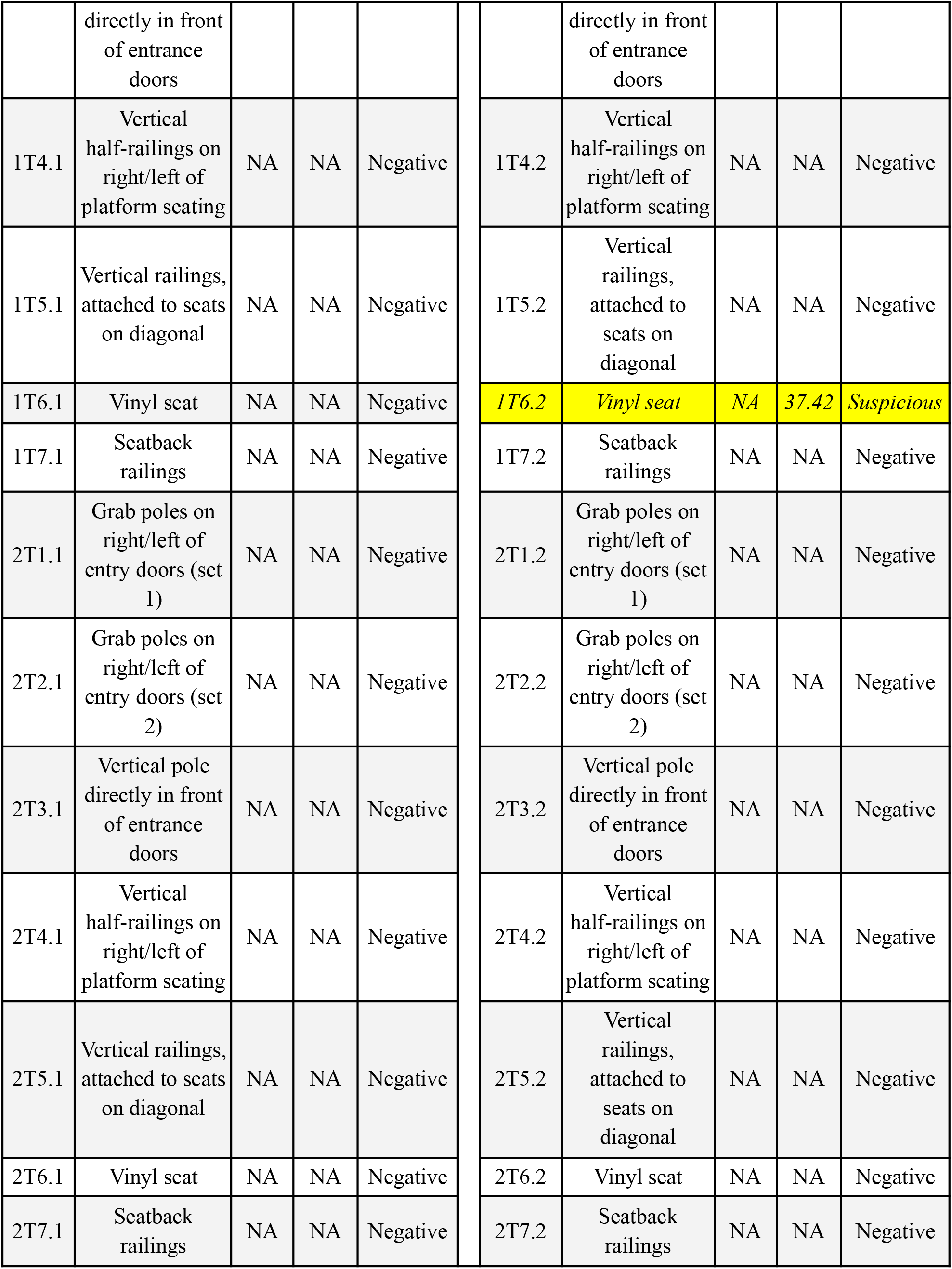
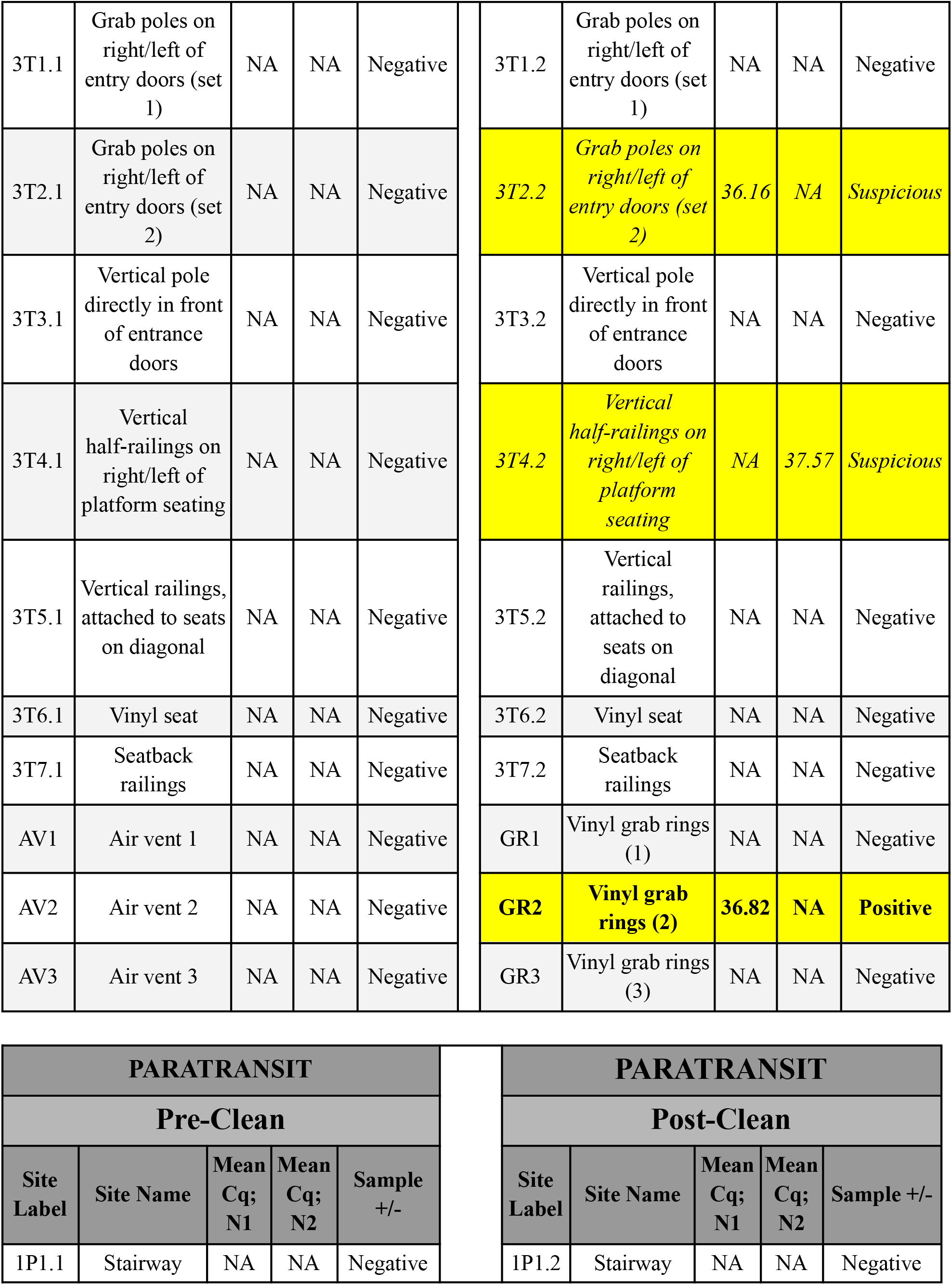
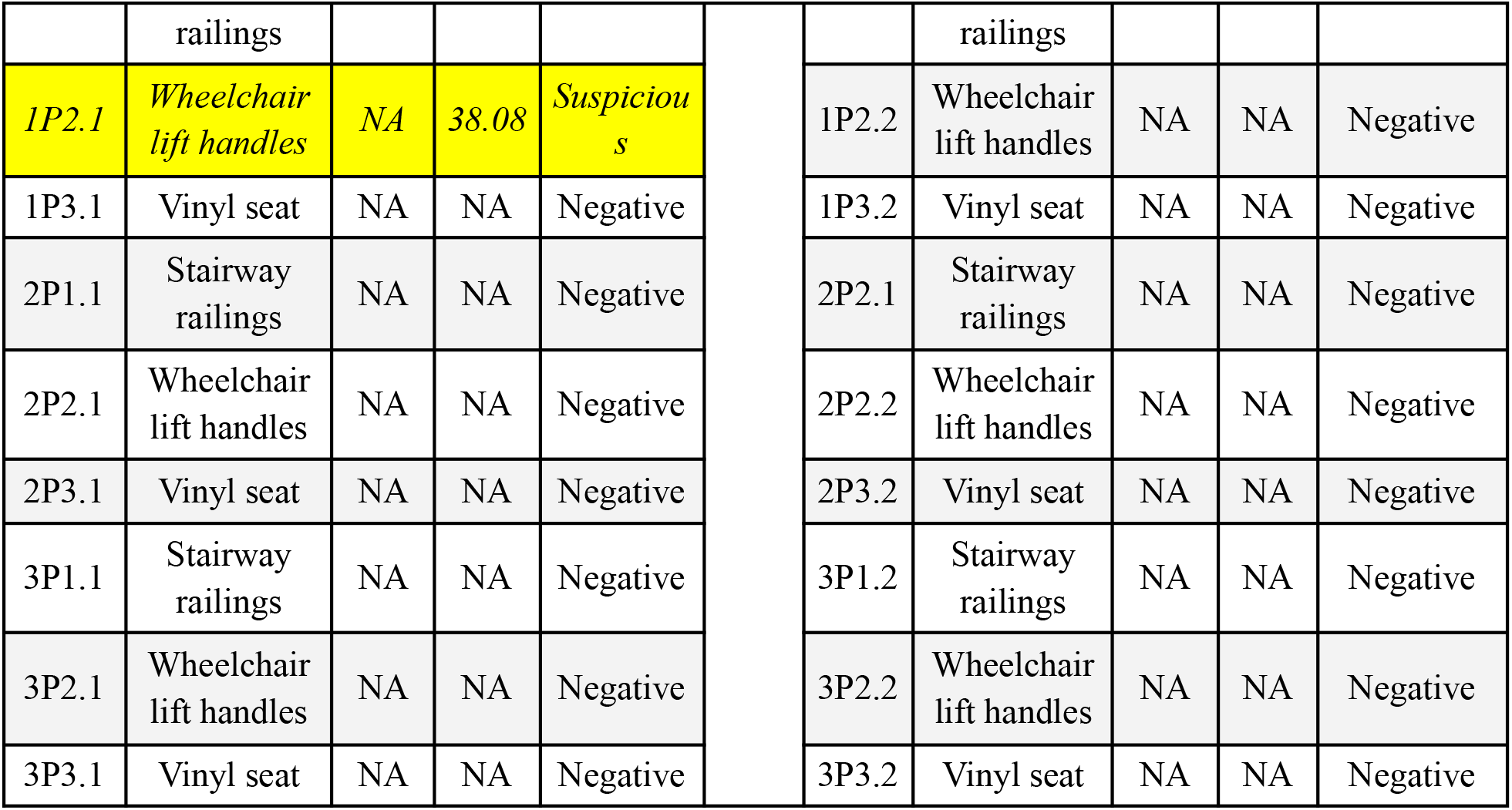
Mean Cq values for each sample collected in Phase III of the experiment. Samples were repeated in triplicate, and the mean of each Cq is reported for both N1 and N2 primers. Cq values highlighted in yellow and italicized are those that were considered suspicious but not positive (only one well reporting positive Cq values), and highlighted in yellow and bolded are those that were considered positive (at least two of three wells reporting positive Cq values).

**Supplementary Table 3.**
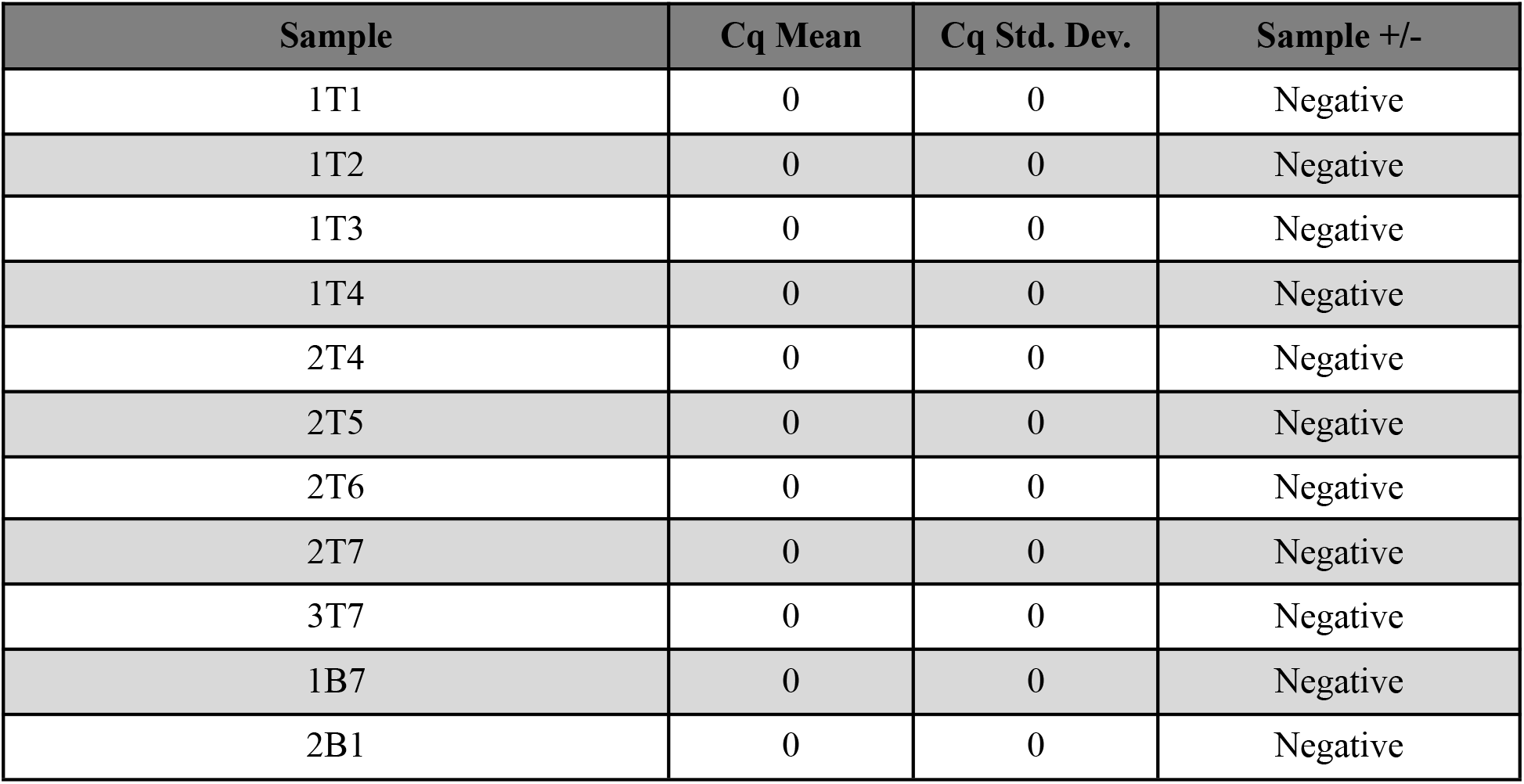
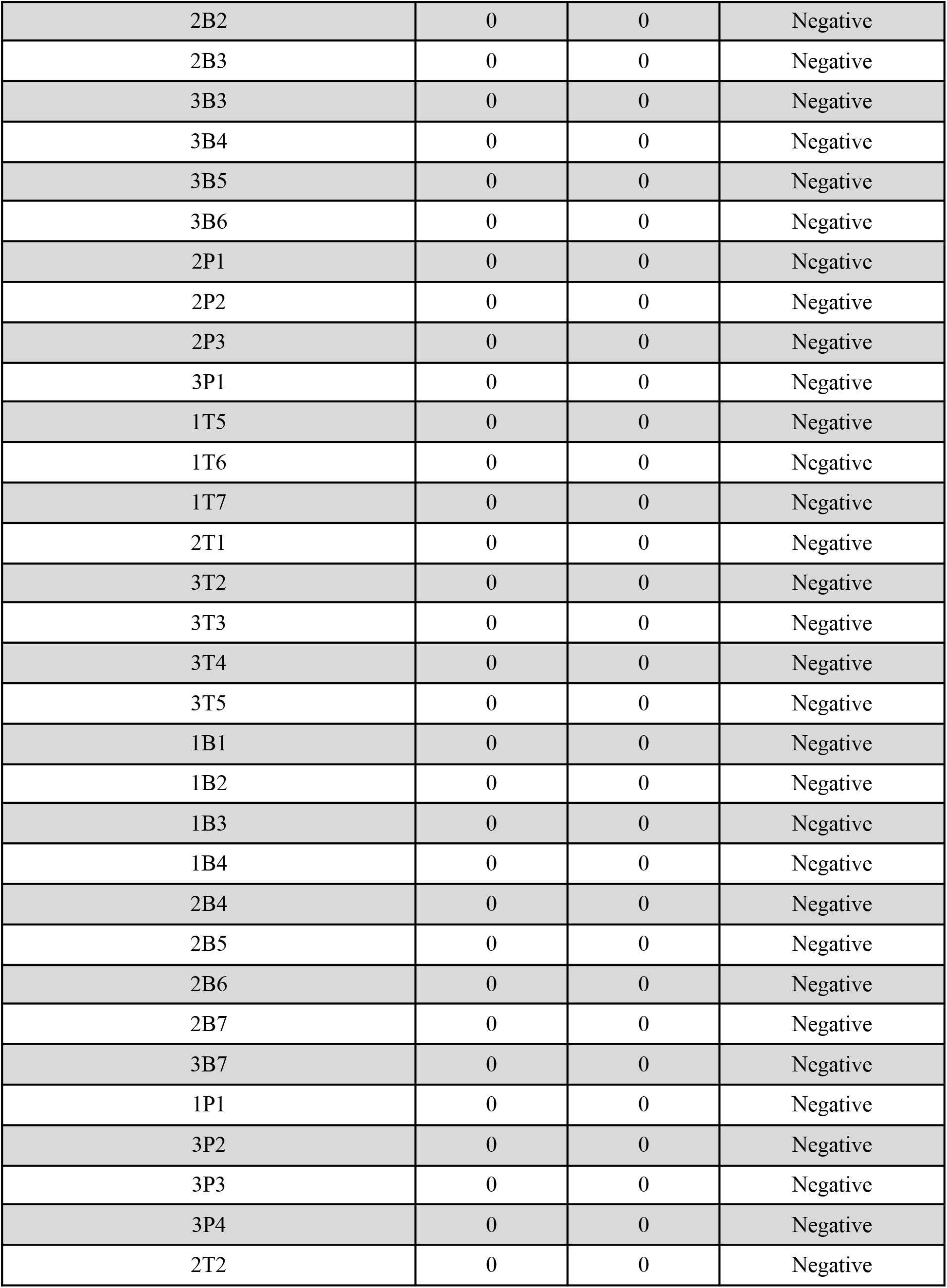
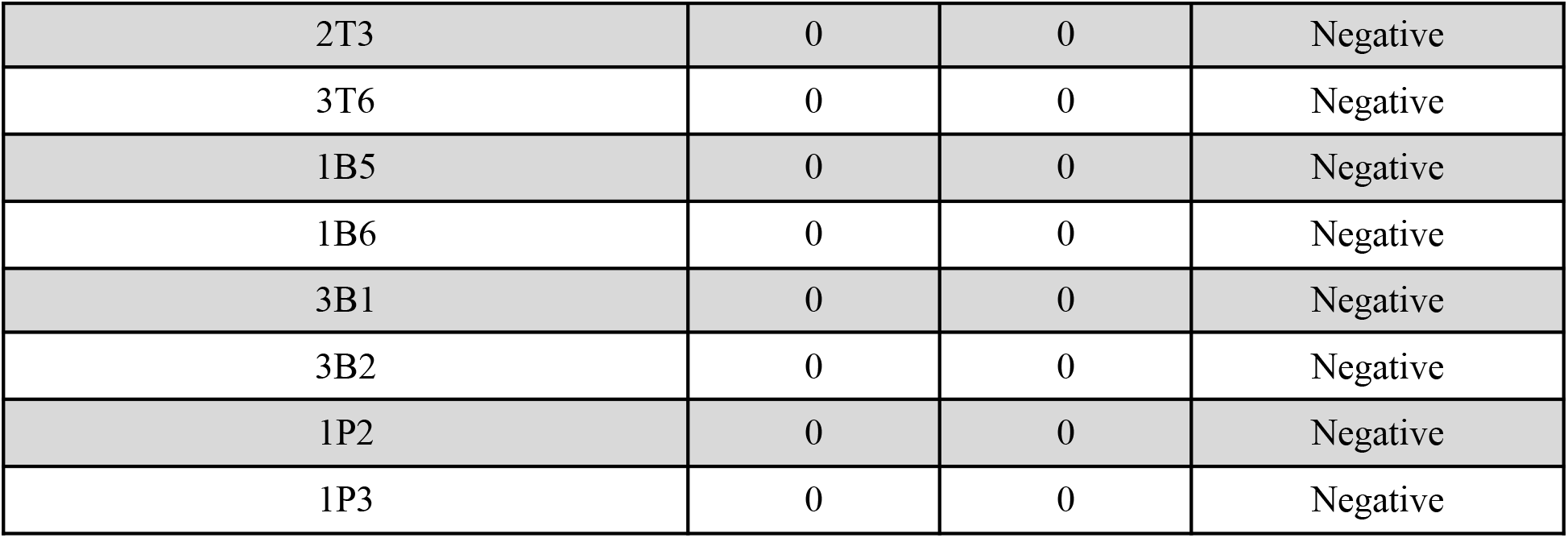
None of the public transit vehicle samples collected during Phase II of this experiment showed any signal in a qPCR analysis using IDT’s N2 primer/probe set.

## References

1. Centers for Disease Control and Prevention. (2020, September 11). Coronavirus Disease 2019 (COVID-19) – Prevention & Treatment.

2. “WHO Coronavirus Disease (COVID-19) Dashboard.” World Health Organization, World Health Organization, 5 Feb. 2021,covid19.who.int/ãgclid=CjwKCAiAgc-ABhA7EiwAjev-jx_4qSGO_9EgN-lS8IJG0DSf4 MOGmo0OrecRv9ARZZBGrW8mHa-dWhoCC_0QAvD_BwE.

3. Vellingiri, B., Jayaramayya, K., Iyer, M., Narayanasamy, A., Govindasamy, V., Giridharan, B., Ganesan, S., Venugopal, A., Venkatesan, D., Ganesan, H., Rajagopalan, K., Rahman, P. K. S. M., Cho, S.-G., Kumar, N. S., & Subramaniam, M. D. (2020). COVID-19: A promising cure for the global panic. Science of The Total Environment, 725, 138277. https://doi.org/10.1016/j.scitotenv.2020.138277

4. Greenhalgh, T., Jimenez, J., Prather, K., Tufecki, Z., Fisman, D., Schooley, R. (2021). Ten scientific reasons in support of airborne transmission of SARS-CoV-2. The Lancet. https://doi.org/10.1016/S0140-6736(21)00869-2

5. Kasloff, S.B., Leung, A., Strong, J.E. et al. Stability of SARS-CoV-2 on critical personal protective equipment. Sci Rep 11, 984 (2021). https://doi.org/10.1038/s41598-020-80098-3

6. van Doremalen, N., Bushmaker, T., Morris, D. H., Holbrook, M. G., Gamble, A., Williamson, B. N., Tamin, A., Harcourt, J. L., Thornburg, N. J., Gerber, S. I., Lloyd-Smith, J. O., de Wit, E., & Munster, V. J. (2020). Aerosol and Surface Stability of SARS-CoV-2 as Compared with SARS-CoV-1. New England Journal of Medicine, 382(16), 1564–1567. https://doi.org/10.1056/nejmc2004973

7. Riddell, S., Goldie, S., Hill, A. et al. The effect of temperature on persistence of SARS-CoV-2 on common surfaces. Virol J 17, 145 (2020). https://doi.org/10.1186/s12985-020-01418-7

8. “Annual Estimates of the Resident Population for Incorporated Places of 50,000 or More, Ranked by July 1, 2019 Population: April 1, 2010 to July 1, 2019”. United States Census Bureau, Population Division. Retrieved May 21, 2020.

9. NinerNotice: Coronavirus Update - March 12. https://emergency.uncc.edu/health-advisories/coronavirus-information/coronavirus-campus-updates

10. Message from the Chancellor - March 17. https://emergency.uncc.edu/health-advisories/coronavirus-information/coronavirus-campus-updates

11. City of Charlotte. (2020, September 13). City of Charlotte COVID-19 Updates. https://charlottenc.gov/covid19/Pages/default.aspx

12. Charlotte Area Transportation Services. (2020, September 4). Social Distancing While Riding. City of Charlotte. https://charlottenc.gov/newsroom/releases/Pages/RA-200409a.aspx

13. “Phase 3 FAQs (Executive Order 169).” NC.gov, www.nc.gov/covid-19/staying-ahead-curve/phase-3-faqs-executive-order-169.

14. “Social Distancing While Riding.” Charlotte NC.gov, charlottenc.gov/newsroom/releases/Pages/RA-200409a.aspx.

15. “CATS Covid Health & Safety.” >City of Charlotte Government, charlottenc.gov/cats/Pages/Covid-Efforts.aspx.

16. “CDC 2019-Novel Coronavirus (2019-NCoV) Real-Time RT-PCR Diagnostic Panel.” FDA, CDC, 1 Dec. 2020, www.fda.gov/media/134922/.

17. Wölfel R, Corman VM, Guggemos W, Seilmaier M, Zange S, Müller MA, Niemeyer D, Jones TC, Vollmar P, Rothe C, Hoelscher M, Bleicker T, Brünink S, Schneider J, Ehmann R, Zwirglmaier K, Drosten C, Wendtner C. Virological assessment of hospitalized patients with COVID-2019. Nature. 2020 May;581(7809):465–469. doi:10.1038/s41586-020-2196-x. Epub 2020 Apr 1. Erratum in: Nature. 2020 Dec;588(7839):E35. PMID: 32235945.

18. Axess 35’ Low Floor Bus Schematic. www.eldorado-ca.com/heavy-duty-bus. Accessed 26 Jan. 2021.

19. Siemens S70 Schematic. www.theurbanist.org/2016/09/19/link-2-0-sound-transit-capital-committee-siemens-lrv-procurement/2000px-s70_lrv_drawing-svg/. Accessed 31 Jan. 2021.

20. Diamond VIP 2200 Schematic. Digital image. Telin Transportation. Telin Transportation Group, 2021. Web. 26 Jan. 2021.

21. Falzone, L., Musso, N., Gattuso, G., Bongiorno, D., Palermo, C.I., Scalia, G. Stefani, S. (2020). Sensitivity assessment of droplet digital PCR for SARS-CoV-2 detection. International Journal of Molecular Medicine, 46, 957–964. https://doi.org/10.3892/ijmm.2020.4673

22. The MetaSUB International Consortium., Mason, C., Afshinnekoo, E. et al. The Metagenomics and Metadesign of the Subways and Urban Biomes (MetaSUB) International Consortium inaugural meeting report. Microbiome 4, 24 (2016). https://doi.org/10.1186/s40168-016-0168-z

