## Supplementary Tables for "Environmental Screening for Surface SARS-CoV-2 Contamination in Urban High-Touch Areas"

**Supplementary Information**

| | Site-Specific Details | Phase II Labels | Phase III Labels |
|---|---|---|---|
| **Paratransit Samples** | Entryway stair railings; stainless steel | P1 | P1.1; P1.2 |
| | Wheelchair lift handles; rubberized | P2 | P2.1; P2.2 |
| | Seat material; vinyl | P3 | P3.1; P3.2 |
| **Train Samples** | Grab rails on right/left of entry doors; set 1; stainless steel | T1 | T1.1; T1.2 |
| | Grab rails on right/left of entry doors; set 2; stainless steel | T2 | T2.1; T2.2 |
| | Vertical grab pole directly in front of entrance doors; stainless steel | T3 | T3.1; T3.2 |
| | Vertical half-railings on right/left of platform seating; stainless steel | T4 | T4.1; T4.2 |
| | Vertical railings; attached to seats on diagonal of platform; stainless steel | T5 | T5.1; T5.2 |
| | Seat material; vinyl | T6 | T6.1; T6.2 |
| | Seatback railings; front/back of train; stainless steel | T7 | T7.1; T7.2 |
| | Air ventilation grate (samples were collected BEFORE sanitization) | -- | AV1, 2, 3 |
| | Grab rings attached to ceiling (samples were collected AFTER sanitization ) | -- | GR1, 2, 3 |
| **Bus Samples** | Ticket kiosk buttons; polyester | B1 | B1.1; B1.2 |
| | Entryway stair railings; stainless steel | B2 | B2.1; B2.2 |
| | Luggage area guard rail; stainless steel | B3 | B3.1; B3.2 |
| | Arm rest; polyester | B4 | B4.1; B4.2 |
| | Stop request lines; rubberized | B5 | B5.1; B5.2 |
| | Ceiling grab rail; stainless steel | B6 | B6.1; B6.2 |
| | Acrylic sneeze guard | B7 | B7.1; B7.2 |
| | Cloth grab ring handles (samples were collected BEFORE sanitization) | -- | H1, 2, 3 |
| **Personal Samples** | Personnel 1 | 1 | 1 |
| | Personnel 2 | 2 | 2 |
| | Personnel 3 | 3 | 3 |

| | | | |
|---|---|---|---|
| | Personnel 4 | 4 | 4 |

**Supplementary Table 1.** Sampling locations with site-specific metadata. Samples collected in Phase II were sampled prior to vehicle sanitization, and samples collected in Phase III were collected before (denoted by a .1) and after (denoted by a .2) vehicle sanitization.

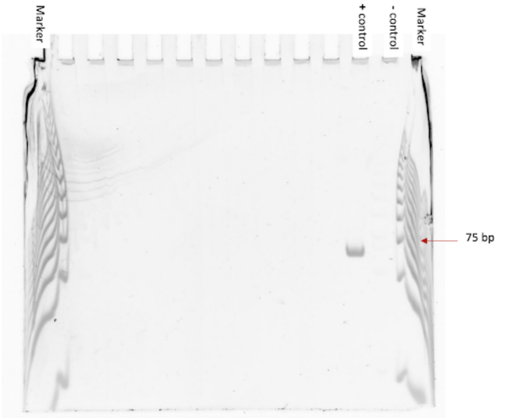


**Supplementary Figure 1.** Example gel image depicting all components indicative of a successful experiment. In this image, the first and last lanes contain the 25 base pair ladder, the 75bp band is indicated. The 2019-nCoV's specific N1 primer produces a PCR product that is 72bp in length, and the 2019-nCov's specific N2 primer produces a PCR product that is 67bp in length. The second to last lane containing the negative control is completely blank, and the positive control shows a band around the 75 base pair mark. A band here indicates a positive result; in the case of our experiment, this would indicate that there is presence of SARS-CoV-2 viral RNA.

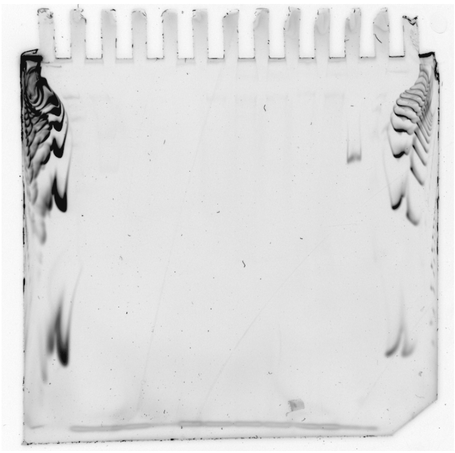

M 1T1 1T2 1T3 1T4 1T5 1T6 1T7 2T1 2T2 2T3 + - M

**Supplementary Figure 2.** Plate 1; row A (all gels for public transit vehicles in Phase II look like this)

| **BUSES** | | | | | **BUSES** | | | | |
|---|---|---|---|---|---|---|---|---|---|
| **Pre-Clean** | | | | | **Post-Clean** | | | | |
| **Site Label** | **Site Name** | **Mean Cq; N1** | **Mean Cq; N2** | **Sample +/-** | **Site Label** | **Site Name** | **Mean Cq; N1** | **Mean Cq; N2** | **Sample +/-** |
| 1B1.1 | Ticket kiosk buttons | NA | NA | Negative | 1B1.2 | Ticket kiosk buttons | NA | NA | Negative |
| 1B2.1 | Entryway stair rail | NA | NA | Negative | 1B2.2 | Entryway stair rail | NA | NA | Negative |
| 1B3.1 | Luggage area guard rail | NA | NA | Negative | 1B3.2 | Luggage area guard rail | NA | NA | Negative |
| 1B4.1 | Horizontal grab rail | NA | NA | Negative | 1B4.2 | Horizontal grab rail | NA | NA | Negative |
| *1B5.1* | *Stop request lines* | *NA* | *37.31* | *Suspicious* | 1B5.2 | Stop request lines | NA | NA | Negative |
| 1B6.1 | Seatback railings | NA | NA | Negative | 1B6.2 | Seatback railings | NA | NA | Negative |
| 1B7.1 | Sneeze guard | NA | NA | Negative | 1B7.2 | Sneeze guard | NA | NA | Negative |
| 2B1.1 | Ticket kiosk buttons | NA | NA | Negative | 2B1.2 | Ticket kiosk buttons | NA | NA | Negative |
| 2B2.1 | Entryway stair rail | NA | NA | Negative | 2B2.2 | Entryway stair rail | NA | NA | Negative |
| 2B3.1 | Luggage area guard rail | NA | NA | Negative | 2B3.2 | Luggage area guard rail | NA | NA | Negative |
| 2B4.1 | Horizontal grab rail | NA | NA | Negative | 2B4.2 | Horizontal grab rail | NA | NA | Negative |
| **2B5.1** | **Stop request lines** | **NA** | **37.74** | **Positive** | 2B5.2 | Stop request lines | NA | NA | Negative |
| 2B6.1 | Seatback | NA | NA | Negative | 2B6.2 | Seatback railings | NA | NA | Negative |

| | | | | | | | | | |
|---|---|---|---|---|---|---|---|---|---|
| | railings | | | | | | | | |
| 2B7.1 | Sneeze guard | NA | NA | Negative | 2B7.2 | Sneeze guard | NA | NA | Negative |
| 3B1.1 | Ticket kiosk buttons | NA | NA | Negative | 3B1.2 | Ticket kiosk buttons | NA | NA | Negative |
| 3B2.1 | Entryway stair rail | NA | NA | Negative | 3B2.2 | Entryway stair rail | NA | NA | Negative |
| 3B3.1 | Luggage area guard rail | NA | NA | Negative | 3B3.2 | Luggage area guard rail | NA | NA | Negative |
| 3B4.1 | Horizontal grab rail | NA | NA | Negative | 3B4.2 | Horizontal grab rail | NA | NA | Negative |
| 3B5.1 | Stop request lines | NA | NA | Negative | 3B5.2 | Stop request lines | NA | NA | Negative |
| 3B6.1 | Seatback railings | NA | NA | Negative | 3B6.2 | Seatback railings | NA | NA | Negative |
| 3B7.1 | Sneeze guard | NA | NA | Negative | 3B7.2 | Sneeze guard | NA | NA | Negative |
| H1 | Cloth grab ring 1 | NA | NA | Negative | | | | | |
| H2 | Cloth grab ring 2 | NA | NA | Negative | | | | | |
| H3 | Cloth grab ring 3 | NA | NA | Negative | | | | | |

| **TRAINS** | | | | | **TRAINS** | | | | |
|---|---|---|---|---|---|---|---|---|---|
| **Pre-Clean** | | | | | **Post-Clean** | | | | |
| **Site Label** | **Site Name** | **Mean Cq; N1** | **Mean Cq; N2** | **Sample +/-** | **Site Label** | **Site Name** | **Mean Cq; N1** | **Mean Cq; N2** | **Sample +/-** |
| 1T1.1 | Grab poles on right/left of entry doors (set 1) | NA | NA | Negative | 1T1.2 | Grab poles on right/left of entry doors (set 1) | NA | NA | Negative |
| 1T2.1 | Grab poles on right/left of entry doors (set 2) | NA | NA | Negative | **1T2.2** | **Grab poles on right/left of entry doors (set 2)** | **35.11** | **38.15** | **Positive** |
| 1T3.1 | Vertical pole | NA | NA | Negative | 1T3.2 | Vertical pole | NA | NA | Negative |

| | | | | | | | | | | |
|---|---|---|---|---|---|---|---|---|---|---|
| | directly in front of entrance doors | | | | | | directly in front of entrance doors | | | |
| 1T4.1 | Vertical half-railings on right/left of platform seating | NA | NA | Negative | | 1T4.2 | Vertical half-railings on right/left of platform seating | NA | NA | Negative |
| 1T5.1 | Vertical railings, attached to seats on diagonal | NA | NA | Negative | | 1T5.2 | Vertical railings, attached to seats on diagonal | NA | NA | Negative |
| 1T6.1 | Vinyl seat | NA | NA | Negative | | *1T6.2* | *Vinyl seat* | *NA* | *37.42* | *Suspicious* |
| 1T7.1 | Seatback railings | NA | NA | Negative | | 1T7.2 | Seatback railings | NA | NA | Negative |
| 2T1.1 | Grab poles on right/left of entry doors (set 1) | NA | NA | Negative | | 2T1.2 | Grab poles on right/left of entry doors (set 1) | NA | NA | Negative |
| 2T2.1 | Grab poles on right/left of entry doors (set 2) | NA | NA | Negative | | 2T2.2 | Grab poles on right/left of entry doors (set 2) | NA | NA | Negative |
| 2T3.1 | Vertical pole directly in front of entrance doors | NA | NA | Negative | | 2T3.2 | Vertical pole directly in front of entrance doors | NA | NA | Negative |
| 2T4.1 | Vertical half-railings on right/left of platform seating | NA | NA | Negative | | 2T4.2 | Vertical half-railings on right/left of platform seating | NA | NA | Negative |
| 2T5.1 | Vertical railings, attached to seats on diagonal | NA | NA | Negative | | 2T5.2 | Vertical railings, attached to seats on diagonal | NA | NA | Negative |
| 2T6.1 | Vinyl seat | NA | NA | Negative | | 2T6.2 | Vinyl seat | NA | NA | Negative |
| 2T7.1 | Seatback railings | NA | NA | Negative | | 2T7.2 | Seatback railings | NA | NA | Negative |

| | | | | | | | | | |
|---|---|---|---|---|---|---|---|---|---|
| 3T1.1 | Grab poles on right/left of entry doors (set 1) | NA | NA | Negative | 3T1.2 | Grab poles on right/left of entry doors (set 1) | NA | NA | Negative |
| 3T2.1 | Grab poles on right/left of entry doors (set 2) | NA | NA | Negative | *3T2.2* | *Grab poles on right/left of entry doors (set 2)* | *36.16* | *NA* | *Suspicious* |
| 3T3.1 | Vertical pole directly in front of entrance doors | NA | NA | Negative | 3T3.2 | Vertical pole directly in front of entrance doors | NA | NA | Negative |
| 3T4.1 | Vertical half-railings on right/left of platform seating | NA | NA | Negative | *3T4.2* | *Vertical half-railings on right/left of platform seating* | *NA* | *37.57* | *Suspicious* |
| 3T5.1 | Vertical railings, attached to seats on diagonal | NA | NA | Negative | 3T5.2 | Vertical railings, attached to seats on diagonal | NA | NA | Negative |
| 3T6.1 | Vinyl seat | NA | NA | Negative | 3T6.2 | Vinyl seat | NA | NA | Negative |
| 3T7.1 | Seatback railings | NA | NA | Negative | 3T7.2 | Seatback railings | NA | NA | Negative |
| AV1 | Air vent 1 | NA | NA | Negative | GR1 | Vinyl grab rings (1) | NA | NA | Negative |
| AV2 | Air vent 2 | NA | NA | Negative | **GR2** | **Vinyl grab rings (2)** | **36.82** | **NA** | **Positive** |
| AV3 | Air vent 3 | NA | NA | Negative | GR3 | Vinyl grab rings (3) | NA | NA | Negative |

| **PARATRANSIT** | | | | | **PARATRANSIT** | | | | |
|---|---|---|---|---|---|---|---|---|---|
| **Pre-Clean** | | | | | **Post-Clean** | | | | |
| **Site Label** | **Site Name** | **Mean Cq; N1** | **Mean Cq; N2** | **Sample +/-** | **Site Label** | **Site Name** | **Mean Cq; N1** | **Mean Cq; N2** | **Sample +/-** |
| 1P1.1 | Stairway | NA | NA | Negative | 1P1.2 | Stairway | NA | NA | Negative |

| | | | | | | | | | | |
|---|---|---|---|---|---|---|---|---|---|---|
| | railings | | | | | | railings | | | |
| *1P2.1* | *Wheelchair lift handles* | *NA* | *38.08* | *Suspicious* | | 1P2.2 | Wheelchair lift handles | NA | NA | Negative |
| 1P3.1 | Vinyl seat | NA | NA | Negative | | 1P3.2 | Vinyl seat | NA | NA | Negative |
| 2P1.1 | Stairway railings | NA | NA | Negative | | 2P2.1 | Stairway railings | NA | NA | Negative |
| 2P2.1 | Wheelchair lift handles | NA | NA | Negative | | 2P2.2 | Wheelchair lift handles | NA | NA | Negative |
| 2P3.1 | Vinyl seat | NA | NA | Negative | | 2P3.2 | Vinyl seat | NA | NA | Negative |
| 3P1.1 | Stairway railings | NA | NA | Negative | | 3P1.2 | Stairway railings | NA | NA | Negative |
| 3P2.1 | Wheelchair lift handles | NA | NA | Negative | | 3P2.2 | Wheelchair lift handles | NA | NA | Negative |
| 3P3.1 | Vinyl seat | NA | NA | Negative | | 3P3.2 | Vinyl seat | NA | NA | Negative |

**Supplementary Table 2.** Mean Cq values for each sample collected in Phase III of the experiment. Samples were repeated in triplicate, and the mean of each Cq is reported for both N1 and N2 primers. Cq values highlighted in yellow and italicized are those that were considered suspicious but not positive (only one well reporting positive Cq values), and highlighted in yellow and bolded are those that were considered positive (at least two of three wells reporting positive Cq values).

| Sample | Cq Mean | Cq Std. Dev. | Sample +/- |
|---|---|---|---|
| 1T1 | 0 | 0 | Negative |
| 1T2 | 0 | 0 | Negative |
| 1T3 | 0 | 0 | Negative |
| 1T4 | 0 | 0 | Negative |
| 2T4 | 0 | 0 | Negative |
| 2T5 | 0 | 0 | Negative |
| 2T6 | 0 | 0 | Negative |
| 2T7 | 0 | 0 | Negative |
| 3T7 | 0 | 0 | Negative |
| 1B7 | 0 | 0 | Negative |
| 2B1 | 0 | 0 | Negative |

| | | | |
|---|---|---|---|
| 2B2 | 0 | 0 | Negative |
| 2B3 | 0 | 0 | Negative |
| 3B3 | 0 | 0 | Negative |
| 3B4 | 0 | 0 | Negative |
| 3B5 | 0 | 0 | Negative |
| 3B6 | 0 | 0 | Negative |
| 2P1 | 0 | 0 | Negative |
| 2P2 | 0 | 0 | Negative |
| 2P3 | 0 | 0 | Negative |
| 3P1 | 0 | 0 | Negative |
| 1T5 | 0 | 0 | Negative |
| 1T6 | 0 | 0 | Negative |
| 1T7 | 0 | 0 | Negative |
| 2T1 | 0 | 0 | Negative |
| 3T2 | 0 | 0 | Negative |
| 3T3 | 0 | 0 | Negative |
| 3T4 | 0 | 0 | Negative |
| 3T5 | 0 | 0 | Negative |
| 1B1 | 0 | 0 | Negative |
| 1B2 | 0 | 0 | Negative |
| 1B3 | 0 | 0 | Negative |
| 1B4 | 0 | 0 | Negative |
| 2B4 | 0 | 0 | Negative |
| 2B5 | 0 | 0 | Negative |
| 2B6 | 0 | 0 | Negative |
| 2B7 | 0 | 0 | Negative |
| 3B7 | 0 | 0 | Negative |
| 1P1 | 0 | 0 | Negative |
| 3P2 | 0 | 0 | Negative |
| 3P3 | 0 | 0 | Negative |
| 3P4 | 0 | 0 | Negative |
| 2T2 | 0 | 0 | Negative |

| | | | |
|---|---|---|---|
| 2T3 | 0 | 0 | Negative |
| 3T6 | 0 | 0 | Negative |
| 1B5 | 0 | 0 | Negative |
| 1B6 | 0 | 0 | Negative |
| 3B1 | 0 | 0 | Negative |
| 3B2 | 0 | 0 | Negative |
| 1P2 | 0 | 0 | Negative |
| 1P3 | 0 | 0 | Negative |

**Supplementary Table 3.** None of the public transit vehicle samples collected during Phase II of this experiment showed any signal in a qPCR analysis using IDT's N2 primer/probe set.
